# Horizon-dependent forecast ranking under structural change: a rolling-origin benchmark for global COVID-19 incidence

**DOI:** 10.64898/2026.03.11.26348121

**Authors:** Michael Marko Sesay, Moise Saleh Wembo

**Author notes:** Department of Mathematics, Pan African University Institute for Basic Sciences, Technology and Innovation (PAUSTI), Juja, Kenya.

## Abstract

Short-horizon epidemic forecasting is difficult when surveillance series are highly nonstationary and affected by structural change and evolving reporting conditions. This study evaluates statistical models for global daily COVID-19 incidence using a rolling-origin benchmark designed to approximate real-time forecasting under such conditions. Using global incidence data from 22 January to 27 July 2020, we compare naive, seasonal naive, drift, ARIMA(log1p), ETS(log1p), and Prophet(log1p) forecasts at horizons *h* ∈ {1, 3, 7, 14} days. Structural phases are identified retrospectively on a variance-stabilized scale and used only to stratify forecast errors. Forecast ranking is strongly horizon-dependent. In the full-sample benchmark, drift performs best at the 1-, 7-, and 14-day horizons, while seasonal naive performs best at 3 days. Among the transformed statistical models, ARIMA(log1p) is competitive at short horizons, whereas ETS(log1p) becomes stronger at 7 and 14 days. Diebold-Mariano tests confirm that several of these differences are statistically meaningful, particularly in favor of drift at short and long horizons and in favor of ETS(log1p) over ARIMA(log1p) at longer horizons. Prophet(log1p) is not competitive in point forecasting and achieves high nominal interval coverage mainly through very wide prediction intervals. Robustness analyses show that the main ranking patterns are broadly stable to alternative segmentation settings, training-window policies, coverage-stabilized subsamples, and alternative target construction based on cumulative confirmed counts. Overall, the results show that simple baselines remain difficult to outperform in epidemic surveillance data and that horizon-specific rolling evaluation is essential for credible forecast comparison under structural change.

**Author summary:** Forecasting infectious disease incidence is difficult when case data change rapidly over time and when reporting systems are still evolving. In this study, I examined how several common statistical forecasting models perform on global daily COVID-19 incidence during the early pandemic. Rather than asking which model is best overall, I focused on whether model ranking changes across forecast horizons and whether those conclusions remain stable under different evaluation choices. I compared simple baselines, including naive, seasonal naive, and drift forecasts, with ARIMA, exponential smoothing, and Prophet models using a rolling-origin benchmark that mimics real-time forecasting. I found that forecast ranking depends strongly on the horizon: drift performed best at 1, 7, and 14 days, while seasonal naive performed best at 3 days. Among the transformed statistical models, ARIMA was more competitive at shorter horizons, whereas exponential smoothing was stronger at longer horizons. I also found that these conclusions remained broadly stable under alternative segmentation settings, training windows, coverage-stabilized subsamples, and target definitions. These results show that simple baselines can remain highly competitive in epidemic surveillance data and that horizon-specific evaluation is essential for fair forecast comparison under structural change.

## Introduction

Accurate short-horizon forecasts of infectious disease incidence support situational awareness, near-term capacity planning, and rapid operational decision-making during epidemics. During the COVID-19 pandemic, forecasting models were widely used by public health agencies and decision-makers to anticipate short-term changes in cases, hospitalizations, and mortality. However, epidemic incidence series are typically highly nonstationary: abrupt changes in level and growth can arise from evolving transmission conditions, immunity, emerging variants, behavioral responses, and changes in surveillance and reporting practices. As a result, forecast accuracy is strongly time-dependent, and model rankings can vary substantially across epidemic phases and forecast horizons [1–3].

Large-scale comparisons from the COVID-19 Forecast Hubs document this variability and show that no single method consistently dominates across time periods or targets. Instead, performance depends on forecast horizon, epidemic phase, and evaluation protocol. In particular, simple statistical baselines often remain highly competitive at short horizons, while more flexible models may perform better under different dynamic conditions [4, 5].

A central challenge is that forecast quality can deteriorate sharply when the data-generating process changes, whether because of epidemiological transitions or reporting artifacts. Consequently, conclusions drawn from a single train–test split can be fragile if the split coincides with an atypical phase. Rolling-origin (walk-forward) backtesting more closely approximates real-time use: models are repeatedly re-estimated using information available up to time *t* and evaluated at multiple horizons beyond *t*, enabling horizon-specific and time-local comparisons of forecast accuracy. Such protocols provide a more reliable assessment of performance under evolving conditions [6–8].

A wide range of approaches has been applied to COVID-19 forecasting, including classical time-series models, machine-learning methods, and mechanistic models. Prior studies consistently report increasing forecast error with horizon and unstable rankings over time. However, fewer studies explicitly examine how structural changes in the observed incidence series interact with horizon-specific model rankings under a rolling-origin framework. In addition, the sensitivity of conclusions to practical evaluation design choices—such as regime definitions, training-window policies, and target construction—remains underexplored, despite its importance for reproducibility and fair benchmarking [9–11].

This study addresses these issues through a transparent statistical benchmark of global daily COVID-19 incidence during the early pandemic. Rather than promoting a single preferred model, we examine how forecast rankings vary across horizons and structural phases, and whether the main conclusions remain stable under alternative evaluation designs. This perspective supports recent calls for more robust and reproducible forecast evaluation in epidemic forecasting.

### Contributions

- **Rolling-origin, horizon-wise benchmark**. We evaluate statistical forecasts at horizons *H* ∈ {1, 3, 7, 14} days using walk-forward backtesting, providing horizon-specific comparisons across commonly used baseline and transformed time-series models.
- **Regime-aware retrospective evaluation**. Structural breaks are detected on a variance-stabilized scale and used only to stratify forecast errors retrospectively, allowing performance to be examined across distinct phases of the observed incidence series.
- **Robustness to evaluation design choices**. We assess the sensitivity of model rankings to alternative segmentation settings, expanding versus sliding training windows, coverage-stabilized subsamples, and alternative target constructions.
- **Empirical insights on horizon-dependent forecasting**. The case study shows that forecast rankings depend strongly on horizon: simple baselines remain highly competitive at short horizons, while transformed statistical models become relatively stronger at longer horizons. The main ranking patterns remain broadly stable across the robustness analyses considered.

The remainder of the paper is organized as follows. Section Material and Methods describes the data and target construction and presents the forecasting models, regime detection procedure, and rolling-origin evaluation protocol. The Section Results report the benchmark and robustness results. Section Discussion discusses the implications and limitations, and Section Conclusion concludes.

## Materials and methods

### Data source and target construction

We analyze a global daily COVID-19 incidence series constructed from the Johns Hopkins University (JHU) CSSE reports distributed via Kaggle (`day_wise.csv`) [12]. The study window spans 22 January 2020 to 27 July 2020, yielding *T* = 188 daily observations. The dataset provides global cumulative totals and daily increments, together with a coverage-related variable (`No. of countries`) recording the number of reporting countries contributing to the aggregate.

Our main forecasting target is the reported daily incidence series

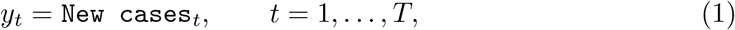

rather than cumulative confirmed counts, since forecasting cumulative totals can yield artificially small errors in a rapidly increasing epidemic. For variance stabilization in selected forecasting models and in retrospective segmentation, we also define

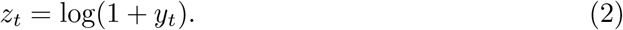

As a robustness check, we consider an alternative target based on first differences of cumulative confirmed counts,

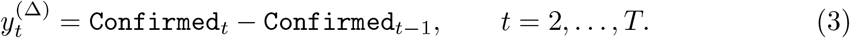

This reconstructed incidence series differs from the reported `New cases` target on a limited subset of dates. Across overlapping dates, the median difference between the reported and reconstructed definitions is zero, and the upper quartile is also zero, indicating that the two series coincide on most days. Nevertheless, a small number of larger mismatches are present, motivating the target-definition robustness analysis reported later.

A second notable feature of the dataset is the monotone increase in `No.of countries`, which rises from 6 to 187 over the study period. We use this variable as a coverage proxy and later exploit it to define coverage-stabilized subsamples, allowing us to assess whether the main forecasting conclusions depend on the early phase of expanding reporting coverage.

### Data diagnostics and interpretation caveat

The global incidence series is strongly nonstationary and highly heteroskedastic. Daily counts range from 0 to 282,756, with a mean of 87,771, a median of 81,114, and a standard deviation of 75,295. The zero rate is low (approximately 0.53%), but both the level and the variability of the series increase sharply over time. Fig 1 shows the raw daily incidence together with its log-transformed companion view, highlighting the pronounced growth phase, evolving volatility, and substantial changes in scale over the early-pandemic period.

**Fig 1.**
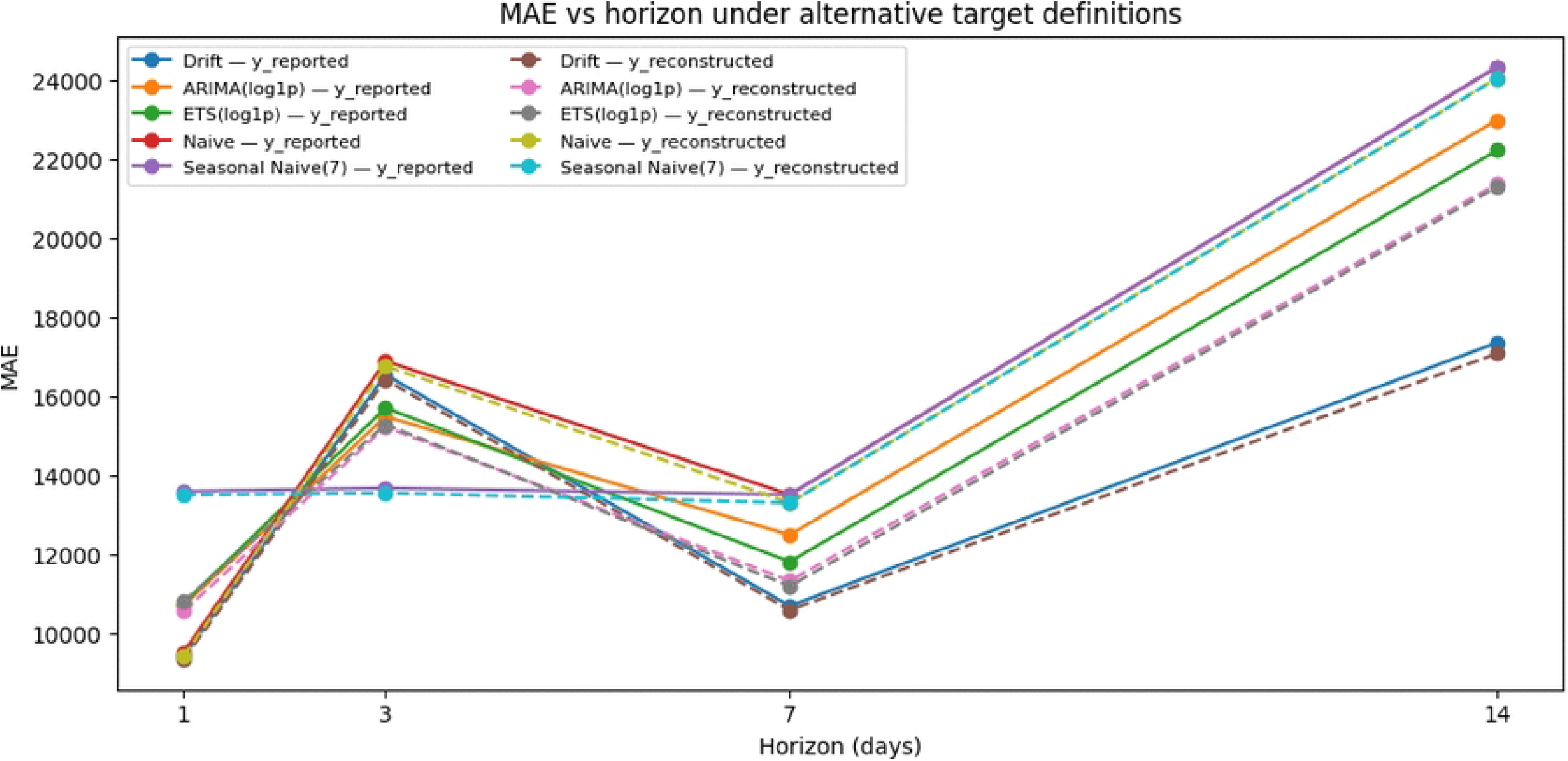
Global daily COVID-19 incidence over the study period. The raw incidence series exhibits strong nonstationarity and increasing variability, while the log-transformed view highlights structural changes in level and growth more clearly.

In addition to the changing level of the series, the coverage proxy grows monotonically and plateaus near the end of the sample (Fig 2). This implies that structural changes in the observed global incidence series reflect not only epidemic dynamics but also evolving reporting coverage and data aggregation. Accordingly, any retrospectively detected regime shifts should be interpreted as changes in the *observed series*, rather than as causal signatures of specific interventions or epidemiological mechanisms.

**Fig 2.**
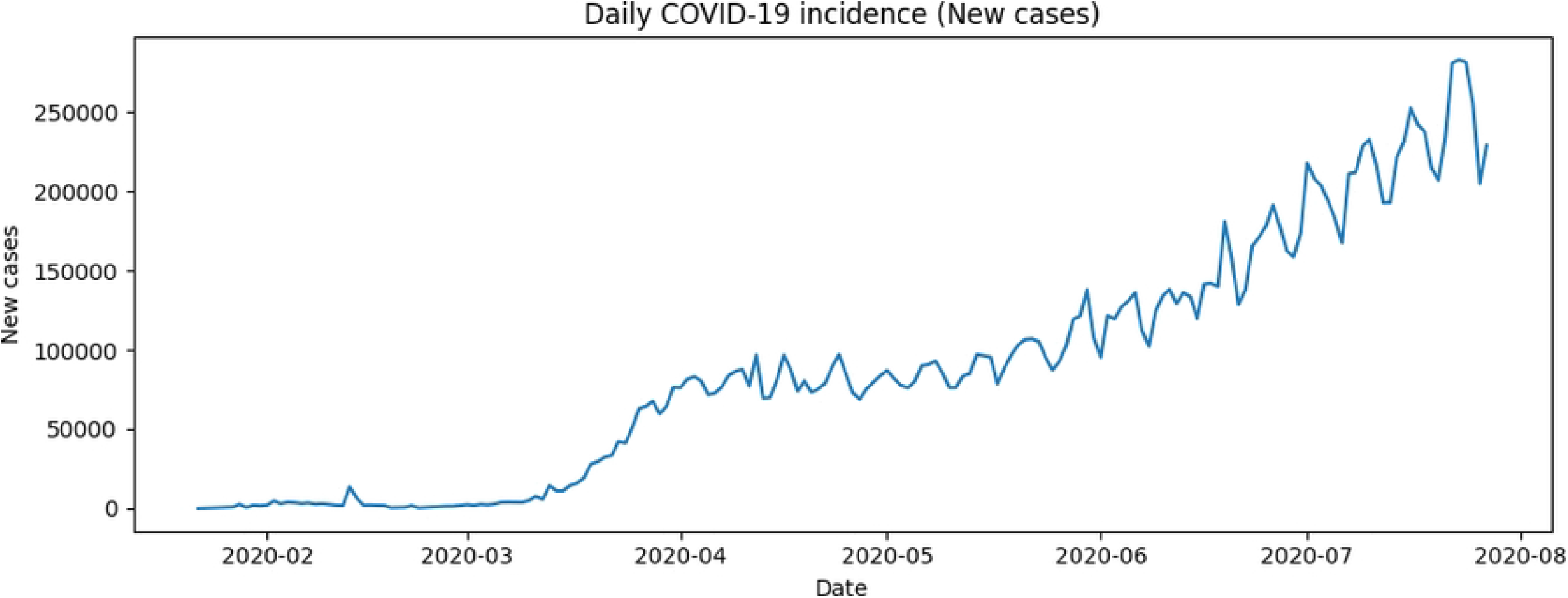
Coverage proxy over time. The number of reporting countries increases monotonically from 6 to 187 and then plateaus near the end of the sample. Vertical reference lines marking the first dates at which the coverage proxy reaches 180, 185, and 187 countries are used later to define coverage-stabilized robustness subsamples.

This interpretation caveat motivates both the rolling-origin evaluation and the robustness analyses. Rolling-origin evaluation is less sensitive than a single train-test split to localized structural features, while the coverage-stabilized and target-definition analyses test whether the main ranking conclusions depend materially on measurement and aggregation effects.

### Forecasting task

We study short- and medium-horizon point forecasting for daily incidence at horizons.

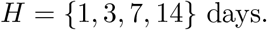

Forecasts are evaluated using a rolling-origin (walk-forward) protocol. Let *t* denote a forecast origin, so that observations up to time *t* − 1 are available for model estimation and the first forecast target is *y*_*t*_. At each origin, we generate multi-step forecasts and evaluate them on the original incidence scale.

Unless stated otherwise, the default estimation policy uses an expanding training window with a minimum training length.

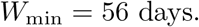

Thus, valid forecast origins are

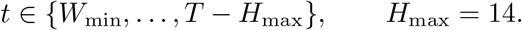

At each origin, models are refit using only information available up to that time, yielding horizon-specific forecast errors aggregated across many origins. All primary accuracy measures—including MAE, RMSE, sMAPE, and MASE—are computed on the original incidence scale, even when models are estimated on the transformed scale log(1 + *y*_*t*_).

The main benchmark is conducted on the full sample using the reported daily incidence target. Additional robustness analyses examine sensitivity to alternative regime definitions, training-window policies, coverage-stabilized subsamples, and the reconstructed incidence target based on cumulative confirmed counts.

1. Daily global incidence (`New cases`).
2. Variance-stabilized incidence, log(1 + *y*_*t*_).

### Methods

This section describes the rolling-origin evaluation protocol, the retrospective regime segmentation procedure, the benchmark forecasting models, the performance metrics, and the robustness analyses.

### Rolling-origin evaluation protocol

To evaluate short-horizon forecast performance under evolving conditions, we use a rolling-origin (walk-forward) backtesting protocol, which more closely approximates real-time forecasting than a single train-test split [10, 13, 14].

Let 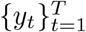 denote the observed daily incidence series. At forecast origin *t*, models are estimated using observations available up to time *t* − 1 and generate forecasts at horizons

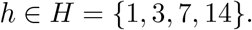

For each horizon *h*, the forecast target is *y*_*t*+*h*_, the expanding training set is

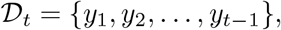

and the forecast is denoted by

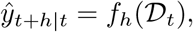

where *f*_*h*_(·) denotes the forecasting model. The corresponding forecast error is

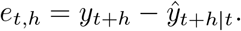

To ensure adequate model estimation, we impose a minimum training length of

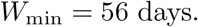

Let *H*_max_ = 14 denote the maximum forecast horizon. Valid forecast origins are therefore

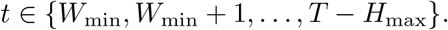

Forecast errors are then aggregated across all valid origins to produce horizon-specific performance summaries.

Fig 3 illustrates the rolling-origin protocol. At each origin, models are trained using only information available at that time and generate forecasts at multiple horizons. This design reduces the sensitivity of model comparisons to any particular train-test split and provides a more realistic assessment of forecast performance under nonstationary conditions [13].

**Fig 3.**
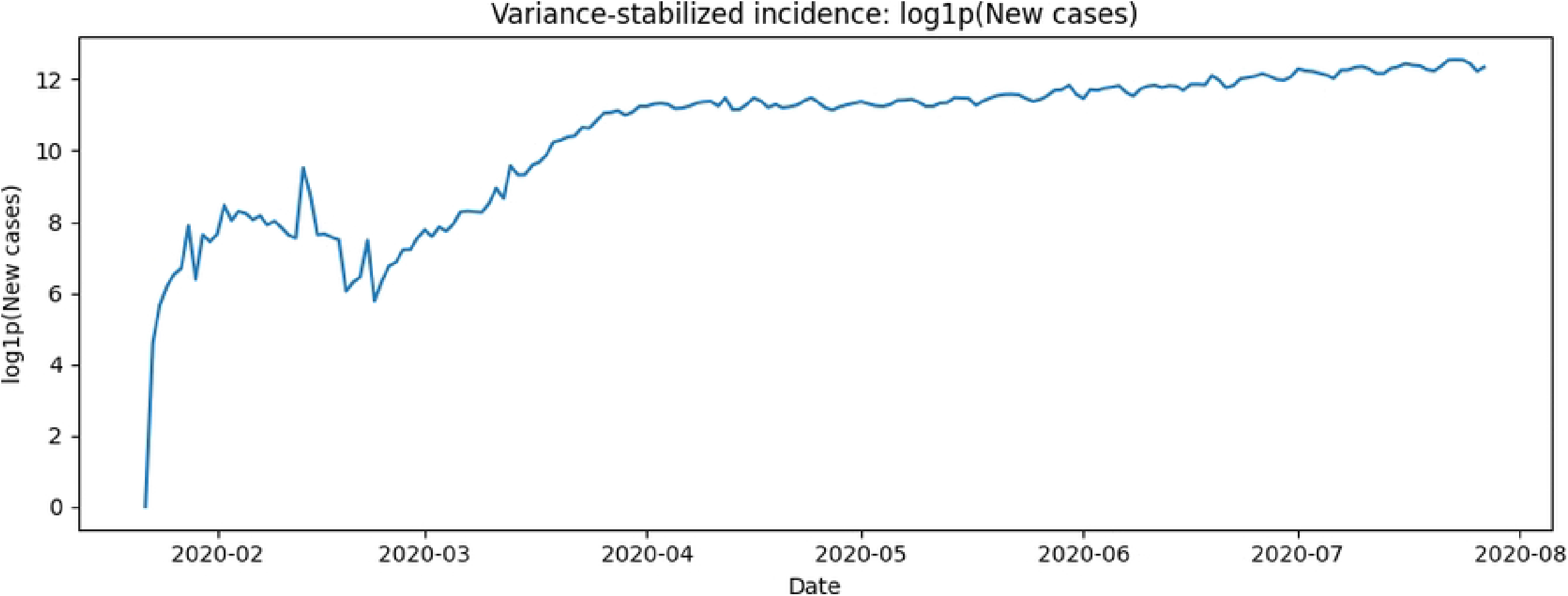
Conceptual illustration of the rolling-origin backtesting protocol. At each forecast origin *t*, models are trained using observations available up to time *t* − 1 and produce forecasts for horizons *h* ∈ {1, 3, 7, 14}. The origin is then advanced in time, yielding multiple evaluation points across the sample.

#### Retrospective regime segmentation

To examine how forecast performance varies across structural phases of the observed incidence series, we perform retrospective regime segmentation on the variance-stabilized series

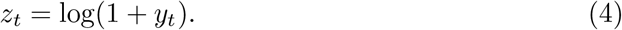

The log transformation reduces heteroskedasticity and stabilizes scale differences over time.

Breakpoints are estimated by minimizing the within-segment sum of squared deviations. For a segment [*i, j*), the cost function is

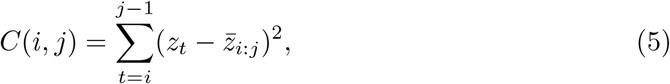

where

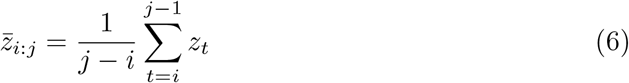

is the segment mean. Given a fixed number of breakpoints *K*, the segmentation problem is

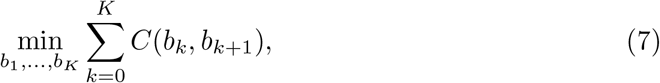

subject to the minimum segment-length constraint

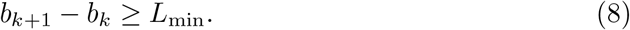

In the baseline specification, we set

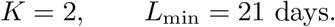

Importantly, regime segmentation is used only for retrospective stratification of forecast errors. The breakpoints are not used for model estimation or forecast generation, thereby avoiding information leakage from future observations into training.

#### Forecasting models

The benchmark includes persistence-based baselines, transformed statistical models, and a reference probabilistic model.

#### Persistence and baseline models

- **Naive**. The naive forecast assumes persistence of the most recent observation:

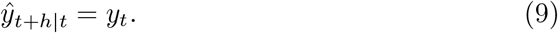
- **Seasonal naive (period** *s* = 7). To capture weekly reporting effects, the seasonal naive forecast repeats the value from the previous week:

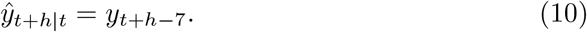
- **Drift**. The drift forecast extrapolates the average historical trend:

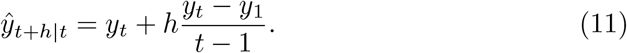

These simple models are standard forecasting benchmarks and often remain highly competitive, particularly at short horizons [13].

#### Transformed statistical models

Two standard time-series models are estimated on the transformed series

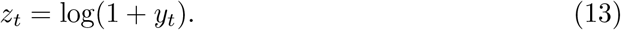

- **ARIMA(log1p)**. Autoregressive integrated moving-average models are fitted to *z*_*t*_, with model order selected by minimizing the Akaike Information Criterion (AIC),

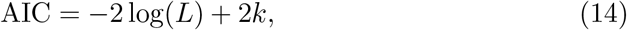

where *L* is the likelihood and *k* is the number of parameters [15].
- **ETS(log1p)**. Exponential smoothing state-space models are fitted to *z*_*t*_ using a small candidate set including additive error with no trend (ANN), additive trend (AAN), and damped additive trend. The final specification is selected by AIC, following standard exponential-smoothing methodology [13]. Forecasts generated on the transformed scale are back-transformed to the original incidence scale using

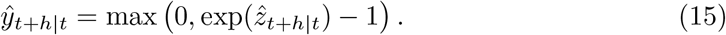

#### Reference probabilistic model

For comparison, we also include Prophet [16], estimated on the transformed series. Prophet decomposes the time series as

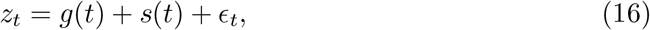

where *g*(*t*) represents a piecewise linear trend and *s*(*t*) captures seasonal effects. In this study, we enable weekly seasonality and disable yearly seasonality due to the short sample span. Prophet is included primarily to examine predictive interval behavior rather than as a strong point-forecast competitor.

### Performance metrics

Forecast accuracy is evaluated using several standard point-forecast metrics.

#### Mean Absolute Error (MAE)

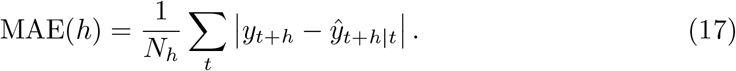

#### Root Mean Squared Error (RMSE)

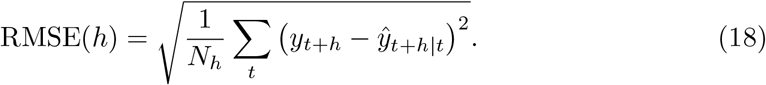

#### Symmetric Mean Absolute Percentage Error (sMAPE)

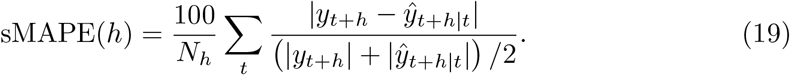

#### Mean Absolute Scaled Error (MASE)

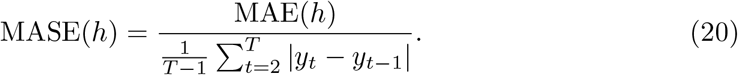

For Prophet, we additionally evaluate interval behavior using empirical coverage probability and mean interval width.

### Formal pairwise forecast comparison

To assess whether pairwise differences in forecast accuracy are statistically significant, we apply the Diebold-Mariano (DM) test [17]. Let *L*(*e*) denote a loss function applied to forecast errors, such as absolute error (AE) or squared error (SE). For two competing models, *A* and *B*, the loss differential is

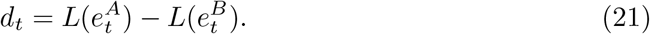

Under the null hypothesis of equal predictive accuracy,

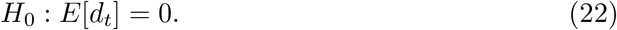

The DM statistic is

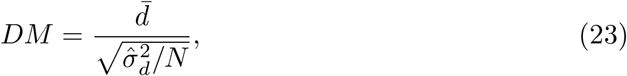

where 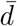 is the sample mean loss differential and 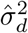 is a heteroskedasticity- and autocorrelation-consistent variance estimate. Significant negative values indicate that model *A* outperforms model *B*, whereas positive values favor model *B*.

### Robustness experiments

To assess the stability of the benchmark conclusions, we conduct four robustness analyses.

- **Segmentation robustness**. Alternative breakpoint configurations and minimum segment lengths are used to assess the stability of retrospective regime definitions.
- **Training-window sensitivity**. Forecast performance is compared under expanding and sliding training windows of different lengths.
- **Coverage-stabilized subsamples**. Subsamples defined by coverage thresholds (e.g., number of reporting countries ≥ 180) are used to assess the influence of early reporting expansion.
- **Target-definition robustness**. The benchmark is repeated using the reconstructed incidence series based on first differences of cumulative confirmed counts.

Together, these experiments assess whether the main ranking conclusions are sensitive to key evaluation design choices.

## Results

### Data overview and structural phases

The global daily COVID-19 incidence series shows pronounced nonstationarity, with large changes in both level and variability over the study period. Fig 4 displays the series together with the baseline retrospective segmentation obtained from the variance-stabilized process *z*_*t*_ = log(1 + *y*_*t*_). Under the baseline specification (*K* = 2, *L*_min_ = 21), two breakpoints are detected, on 13 March 2020 and 28 May 2020, partitioning the sample into three structural phases.

**Fig 4.**
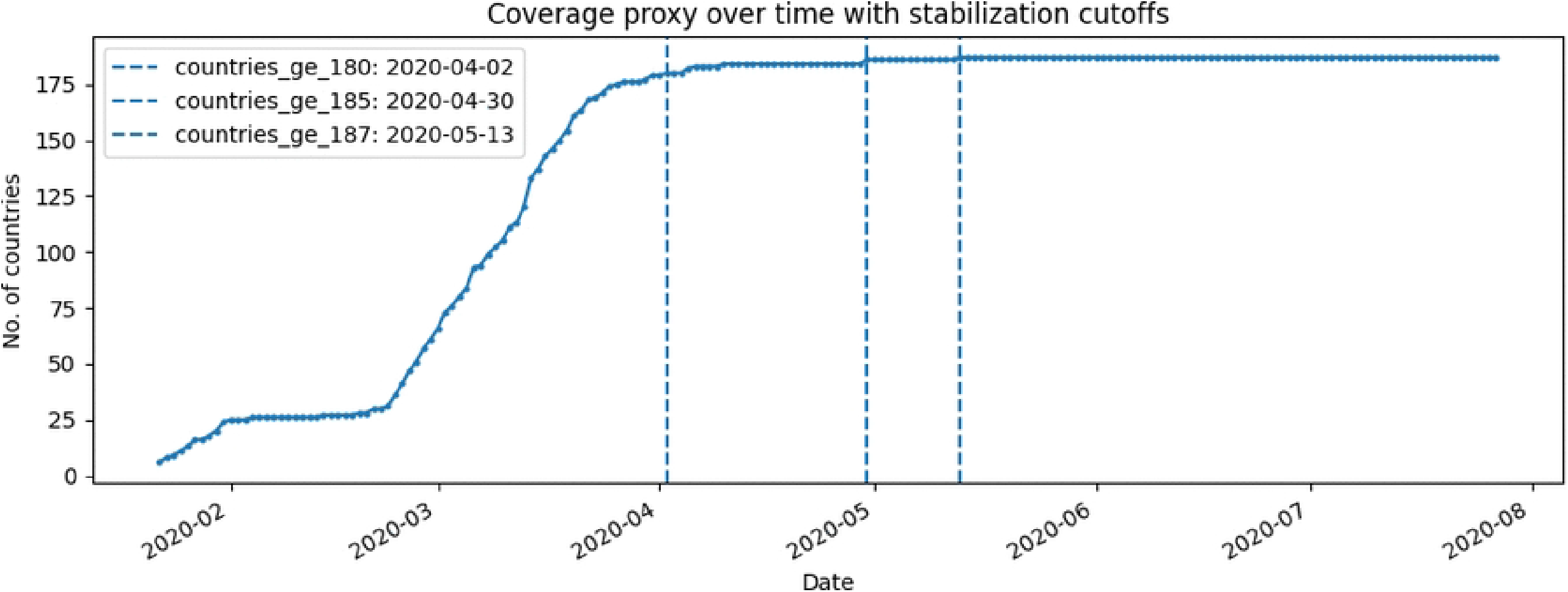
Daily global COVID-19 incidence with detected regime boundaries. Breakpoints are detected on the variance-stabilized series *z*_*t*_ = log(1 + *y*_*t*_) using the baseline segmentation setting (*K* = 2, *L*_min_ = 21). The resulting boundaries occurred on 13 March 2020 and 28 May 2020.

These phases are interpreted as *observed-series regimes* rather than purely epidemiological regimes. In particular, the early part of the sample reflects both rapid growth in incidence and substantial expansion in reporting coverage, as indicated by the monotone increase in the number of contributing countries. Accordingly, the detected breakpoints summarize changes in the observed global aggregate and are used only for retrospective stratification of forecast errors.

Table 2 summarizes the three baseline regimes. Regime 1 (22 January–12 March 2020) corresponds to the initial low-incidence phase, characterized by the smallest average case counts and the lowest reporting coverage. Regime 2 (13 March–27 May 2020) captures the sharp global escalation phase, during which mean daily incidence increases substantially, and the coverage proxy approaches its plateau. Regime 3 (28 May–27 July 2020) represents a sustained high-incidence phase under near-complete coverage, with the largest average daily counts and a comparatively more stable reporting footprint.

**Table 1.** Data summary and target diagnostics. Summary statistics are reported for the main incidence target *y*_*t*_ = `New cases`_*t*_ over the study window 22 January 2020 to 27 July 2020. The final rows summarize data-quality features relevant to the robustness analyses.

| Metric | Value |
| --- | --- |
| Start date | 2020-01-22 |
| End date | 2020-07-27 |
| Number of days | 188 |
| Minimum daily incidence | 0 |
| Median daily incidence | 81,114 |
| Mean daily incidence | 87,771.02 |
| Maximum daily incidence | 282,756 |
| Standard deviation | 75,295.29 |
| Zero rate | 0.0053 |
| Missing rate | 0.0000 |
| Maximum number of countries | 187 |
| Mismatch count: reported vs reconstructed target | 28 |
| Maximum absolute mismatch | 10,034 |

**Table 2.** Baseline regime summary for global daily COVID-19 incidence. Regimes are obtained from retrospective change-point segmentation of *z*_*t*_ = log(1 + *y*_*t*_) with *K* = 2 breakpoints and minimum segment length *L*_min_ = 21 days. Incidence statistics are reported on the original scale.

| Reg. | Start | End | Days | Mean incidence | SD | Mean log1p | Mean countries |
| --- | --- | --- | --- | --- | --- | --- | --- |
| 1 | 2020-01-22 | 2020-03-12 | 51 | 2,569.57 | 2,304.84 | 7.3665 | 42.10 |
| 2 | 2020-03-13 | 2020-05-27 | 76 | 73,517.45 | 24,373.75 | 11.1050 | 178.74 |
| 3 | 2020-05-28 | 2020-07-27 | 61 | 176,763.57 | 49,428.13 | 12.0437 | 187.00 |

Overall, the series exhibits major changes in scale, dispersion, and reporting context. This supports the use of rolling-origin evaluation and motivates the regime-wise and robustness analyses reported below.

### Main rolling-origin benchmark across models and horizons

Table 3 and Fig 5–6 summarize the main rolling-origin benchmark across horizons *h* ∈ {1, 3, 7, 14}. The dominant feature is strong horizon dependence in model ranking.

**Table 3.** Overall rolling-origin benchmark across forecast horizons. All metrics are computed on the original incidence scale. Lower values indicate better point-forecast accuracy.

**(a) Mean Absolute Error (MAE)**
| Model | $h = 1$ | $h = 3$ | $h = 7$ | $h = 14$ |
| --- | --- | --- | --- | --- |
| Naive | 9526.42 | 16903.04 | 13524.58 | 24342.65 |
| Seasonal Naive(7) | 13607.61 | 13687.83 | 13524.58 | 24342.65 |
| Drift | 9424.21 | 16555.44 | 10708.80 | 17367.38 |
| ARIMA(log1p) | 10716.11 | 15497.77 | 12508.35 | 23002.28 |
| ETS(log1p) | 10761.58 | 15718.33 | 11827.27 | 22233.07 |
| Prophet(log1p) | 50307.81 | 56478.30 | 72221.35 | 118411.71 |

**(b) Root Mean Squared Error (RMSE)**
| Model | $h = 1$ | $h = 3$ | $h = 7$ | $h = 14$ |
| --- | --- | --- | --- | --- |
| Naive | 13085.91 | 21122.01 | 17406.54 | 30179.10 |
| Seasonal Naive(7) | 17497.85 | 17577.16 | 17406.54 | 30179.10 |
| Drift | 13047.00 | 20800.02 | 14127.78 | 22185.07 |
| ARIMA(log1p) | 13685.85 | 19405.89 | 16562.68 | 29049.73 |
| ETS(log1p) | 13791.29 | 19713.98 | 15724.02 | 27959.17 |
| Prophet(log1p) | 66782.06 | 76450.46 | 99996.86 | 172869.44 |

**Fig 5.**
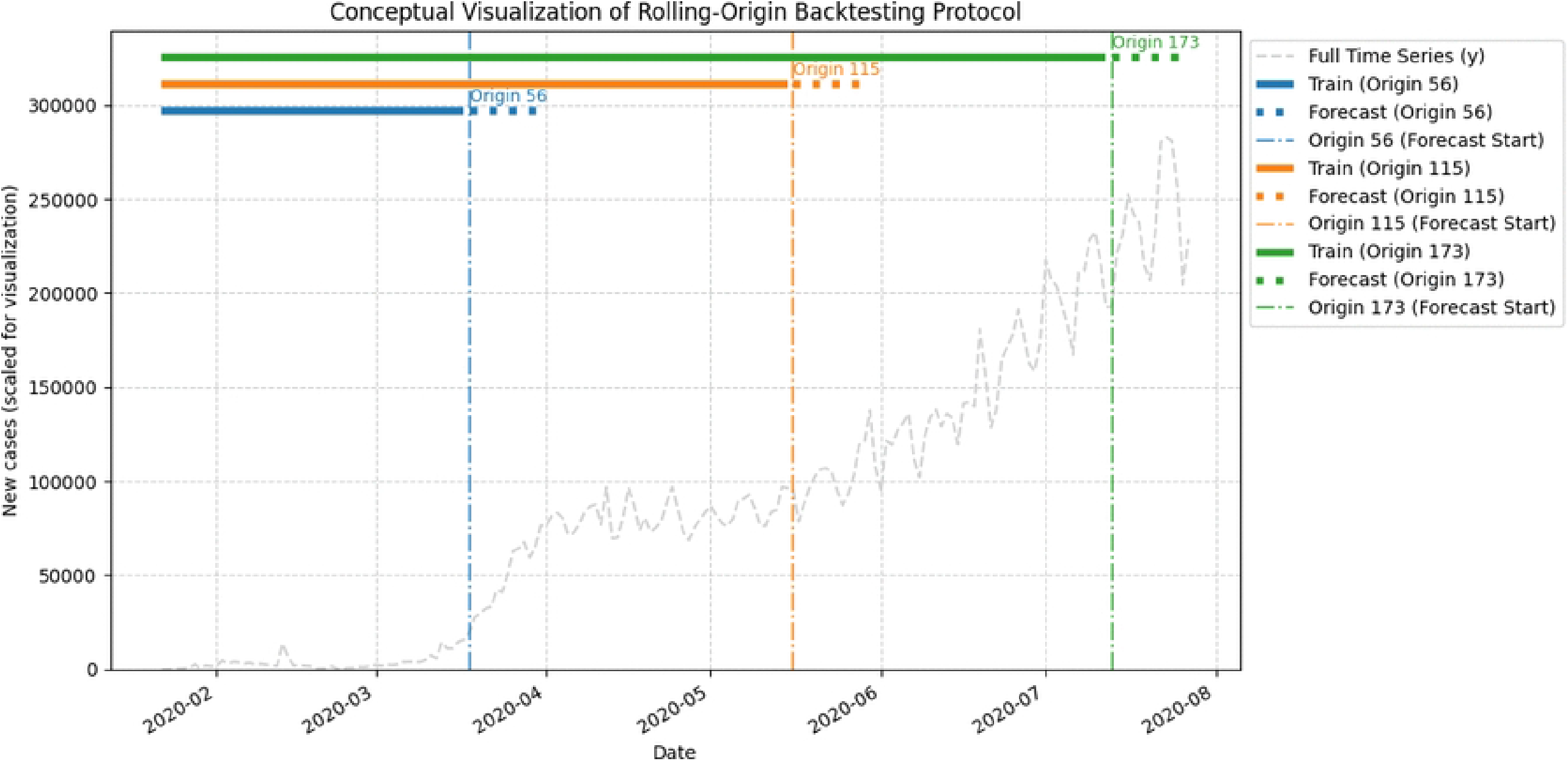
MAE versus forecast horizon for all benchmark models. Forecast ranking varies substantially with horizon. Drift is strongest at 1-, 7-, and 14-day horizons, seasonal naive performs best at 3 days, and Prophet(log1p) is markedly weaker than the other models throughout.

Drift attains the lowest MAE at *h* = 1, *h* = 7, and *h* = 14, showing that a simple trend-extrapolation benchmark remains highly competitive in this global early-pandemic series. At *h* = 3, seasonal naive achieves the lowest MAE, indicating that short weekly recurrence retains predictive value despite the pronounced nonstationary trend in the aggregate series.

ARIMA(log1p) and ETS(log1p) are competitive but not uniformly dominant. Their performance is close at short horizons, with ARIMA(log1p) slightly outperforming ETS(log1p) at *h* = 1 and *h* = 3 in terms of MAE. At longer horizons, however, ETS(log1p) improves relative to ARIMA(log1p), outperforming it at *h* = 7 and *h* = 14. Thus, the relative advantage of these transformed statistical models depends materially on the forecast horizon.

Naive persistence also remains strong at *h* = 1, where it is only slightly worse than drift, but it deteriorates more quickly at longer horizons. This is consistent with the idea that one-step persistence can be effective when the series is locally smooth, whereas longer-horizon prediction benefits more from explicit trend adaptation.

Prophet(log1p) performs poorly throughout. Its MAE and RMSE values are substantially larger than those of the other models at all horizons, indicating that the lightweight specification used here is not competitive for point forecasting in this setting.

Fig 5 visualizes the horizon-wise MAE profiles and makes clear that no single model uniformly dominates. Fig 6 complements this by showing average rank across forecast origins, again highlighting the strong overall performance of drift and the horizon-specific trade-off between ARIMA(log1p) and ETS(log1p).

**Fig 6.**
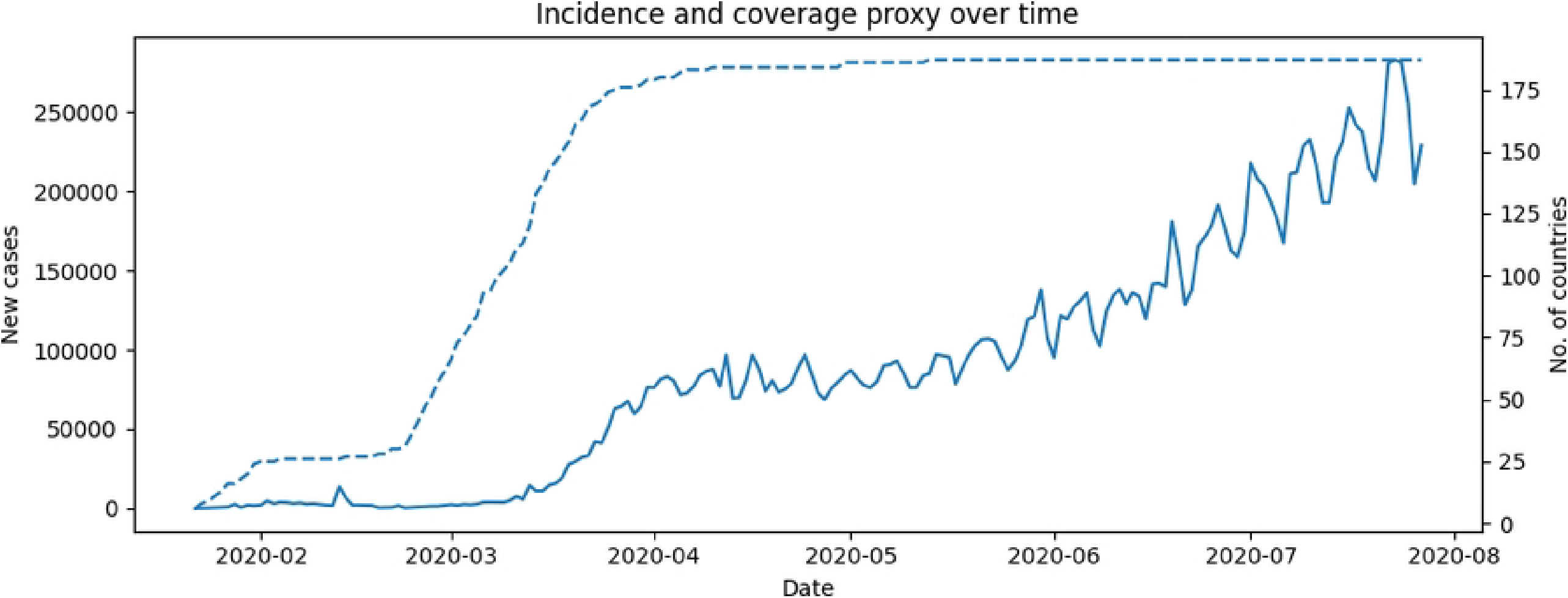
Average rank versus forecast horizon (lower is better). Average rank across rolling forecast origins confirms that model performance is horizon-dependent. Drift is the most consistently competitive benchmark overall, while ARIMA(log1p) and ETS(log1p) remain close and exchange relative advantage across horizons.

Overall, the full-sample benchmark establishes the central empirical result of the paper: forecast ranking is horizon-dependent, and strong simple baselines materially shape the conclusions of comparative forecast evaluation. The statistical significance of the main pairwise differences is assessed in Section (results dm).

### Regime-wise benchmark under structural change

To examine how forecast performance varies across structural phases of the observed incidence series, we stratify rolling-origin forecast errors by the baseline retrospective regimes introduced in Section (results regimes). Regime labels are assigned according to the regime of the last observed point at each forecast origin, so the summaries remain strictly retrospective and do not affect model fitting.

Table 4 and Fig 7 show that horizon dependence persists across regimes, although model ordering changes across phases. In Regime 2 (13 March–27 May 2020), corresponding to the rapid escalation phase under expanding global coverage, drift attains the lowest MAE at *h* = 1 and *h* = 14, while ETS(log1p) performs best at *h* = 7. At *h* = 3, ARIMA(log1p), ETS(log1p), and seasonal naive are close, with seasonal naive achieving the smallest MAE.

**Table 4.** Regime-wise rolling-origin benchmark (MAE). Forecast errors are stratified retrospectively by baseline regime. Lower values indicate better point-forecast accuracy. Prophet(log1p) is omitted for readability and discussed separately in Section (results_prophet).

| Regime | Model | $h = 1$ | $h = 3$ | $h = 7$ | $h = 14$ |
| --- | --- | --- | --- | --- | --- |
| Regime 2 | Naive | 6512.47 | 12545.11 | 11060.81 | 16785.51 |
|  | Drift | 6335.03 | 12313.57 | 9851.80 | 13933.95 |
|  | ARIMA(log1p) | 7324.59 | 11737.08 | 10056.09 | 15774.46 |
|  | ETS(log1p) | 7278.94 | 11751.05 | 9335.63 | 15141.77 |
| Regime 3 | Naive | 14143.53 | 23579.02 | 17298.87 | 35919.53 |
|  | Drift | 14156.57 | 23053.62 | 12021.64 | 22627.10 |
|  | ARIMA(log1p) | 15911.63 | 21258.83 | 16265.00 | 34074.68 |
|  | ETS(log1p) | 16096.70 | 21795.86 | 15644.26 | 33096.34 |

**Fig 7.**
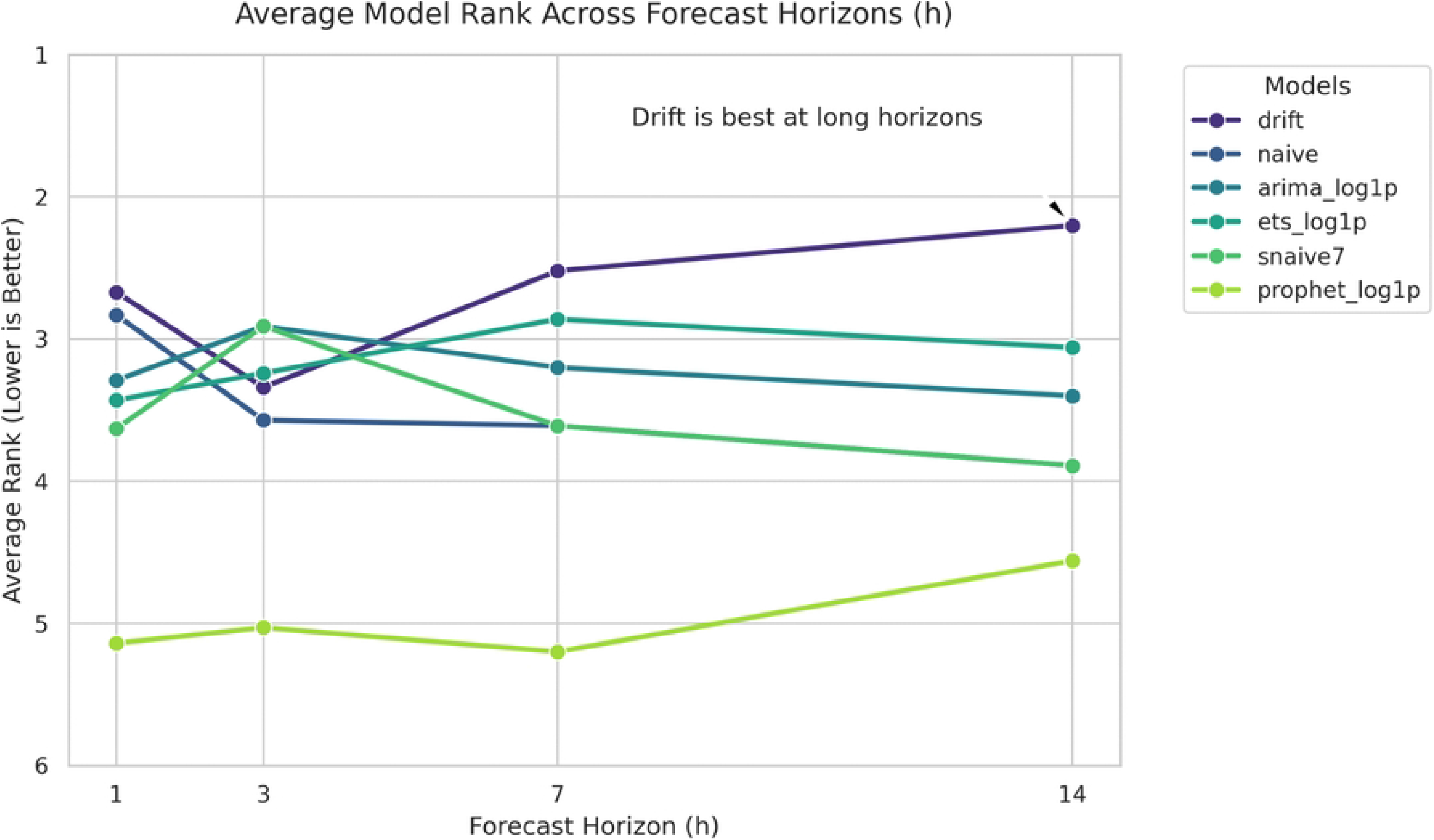
Regime-wise MAE heatmaps across forecast horizons. The relative ordering of models varies across regimes and horizons. Drift and ETS (log1p) remain among the most competitive models, but no single method dominates across all structural phases and horizons.

In Regime 3 (28 May–27 July 2020), corresponding to a sustained high-incidence phase under near-complete coverage, the ranking shifts again. Naive and drift are nearly tied at *h* = 1, with naive slightly better. At *h* = 3, seasonal naive again attains the lowest MAE, while drift is strongest at *h* = 7 and *h* = 14. ETS(log1p) remains competitive in this later phase and improves on naive at *h* = 3, *h* = 7, and *h* = 14, but it does not dominate uniformly.

Two broad patterns emerge. First, the benchmark remains strongly horizon-dependent within each regime, reinforcing the conclusion that model performance cannot be summarized adequately by a single overall winner. Second, drift and ETS (log1p) remain among the most competitive models across both structural phases, although their relative advantage depends on the forecast horizon. This phase sensitivity helps explain why full-sample averages can mask meaningful local differences in forecast behavior.

Prophet (log1p) shows a notable regime-wise contrast. Its point errors are extremely large throughout Regime 2, whereas in Regime 3, its MAE decreases substantially and becomes less extreme relative to the other models, especially at *h* = 14. Even so, Prophet remains uncompetitive overall, and its performance is more informative from the perspective of interval behavior than point accuracy.

Overall, the regime-stratified results support the same central conclusion as the full-sample benchmark: forecast ranking varies across both horizon and structural phase, and simple baselines remain difficult to outperform consistently even when transformed statistical models become more competitive in particular regimes.

**(a)** *h* = 1 **(b)** *h* = 3

**(c)** *h* = 7 **(d)** *h* = 14

### Formal pairwise forecast comparisons

The descriptive benchmark results in Sections (results main benchmark)–(results regimewise) show that forecast ranking varies across horizons and that several differences are relatively small. To assess whether the main pairwise differences are statistically meaningful, we apply Diebold-Mariano (DM) tests based on the absolute-error (AE) loss differential. We focus on three comparisons: drift versus ARIMA(log1p), drift versus ETS(log1p), and ETS(log1p) versus ARIMA(log1p). The AE-based results are reported in Table 5 and summarized in Fig 8.

**Table 5.** Diebold-Mariano test summary based on absolute-error loss. Each row reports the better-performing model for a given pairwise comparison and forecast horizon, together with the two-sided *p*-value. Significance codes: ^∗^*p <* 0.10, ^∗∗^*p <* 0.05, ^∗∗∗^*p <* 0.01. “ns” denotes a non-significant difference.

| Comparison | Horizon | Result |
| --- | --- | --- |
| Drift vs ARIMA(log1p) | $h = 1$ | Drift ($p = 0.0087$)*** |
| Drift vs ARIMA(log1p) | $h = 3$ | ARIMA(log1p) ($p = 0.0526$)* |
| Drift vs ARIMA(log1p) | $h = 7$ | Drift ($p = 0.0337$)** |
| Drift vs ARIMA(log1p) | $h = 14$ | Drift ($p = 0.0299$)** |
| Drift vs ETS(log1p) | $h = 1$ | Drift ($p = 0.0055$)*** |
| Drift vs ETS(log1p) | $h = 3$ | ETS(log1p) ($p = 0.0941$)* |
| Drift vs ETS(log1p) | $h = 7$ | Drift (ns; $p = 0.1695$) |
| Drift vs ETS(log1p) | $h = 14$ | Drift ($p = 0.0486$)** |
| ETS(log1p) vs ARIMA(log1p) | $h = 1$ | ARIMA(log1p) (ns; $p = 0.3579$) |
| ETS(log1p) vs ARIMA(log1p) | $h = 3$ | ARIMA(log1p) ($p = 0.0776$)* |
| ETS(log1p) vs ARIMA(log1p) | $h = 7$ | ETS(log1p) ($p = 0.0157$)** |
| ETS(log1p) vs ARIMA(log1p) | $h = 14$ | ETS(log1p) ($p = 0.0331$)** |

\*

**Fig 8.**
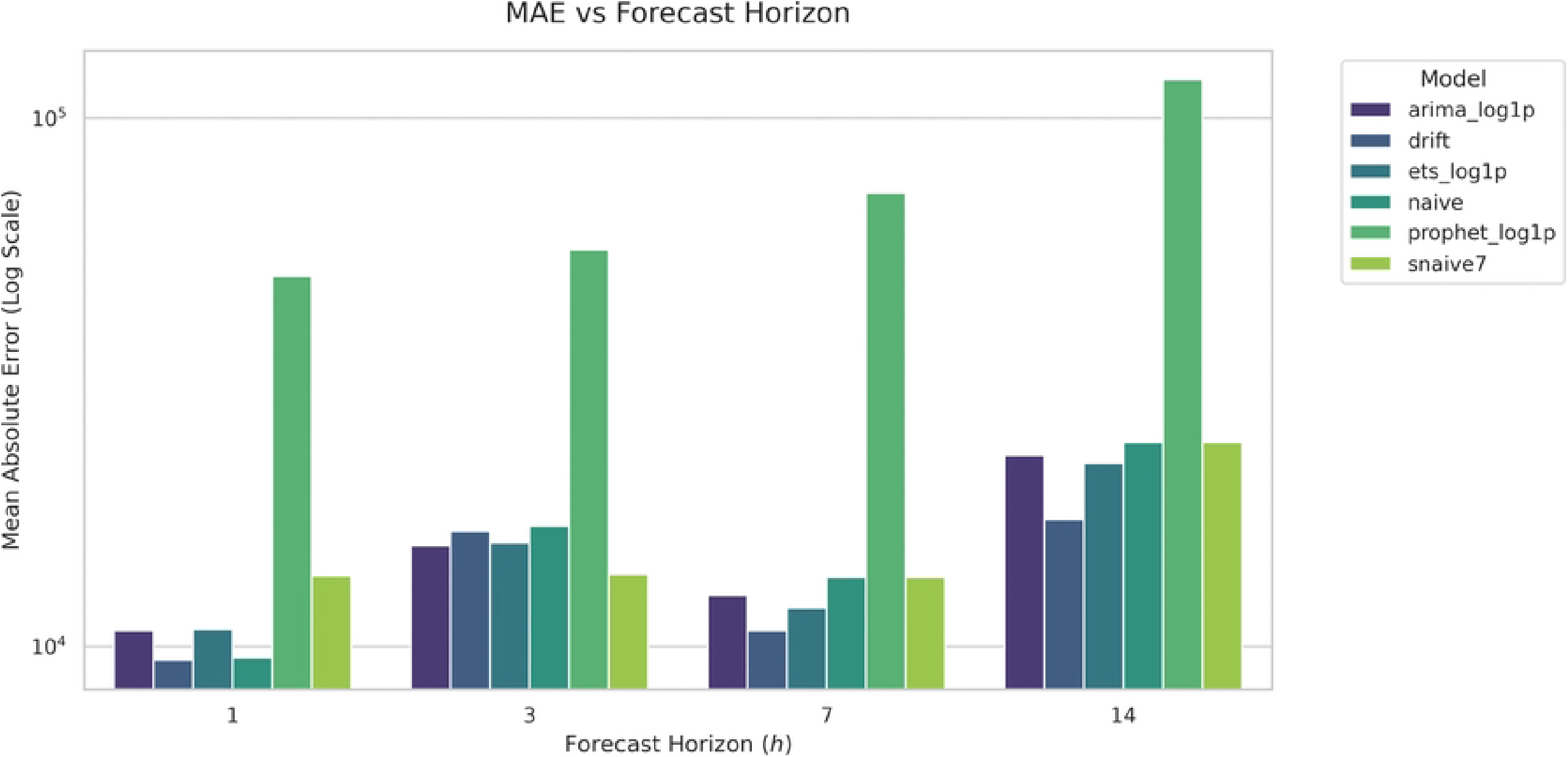
Heatmap summary of Diebold-Mariano comparisons under absolute-error loss. The heatmap reports the better-performing model for each pairwise comparison and forecast horizon, together with significance coding. Drift is significantly stronger than both ARIMA(log1p) and ETS(log1p) at several horizons, while ETS(log1p) significantly outperforms ARIMA(log1p) at 7 and 14 days.

The pairwise tests reinforce the horizon-dependent ranking pattern. Drift significantly outperforms ARIMA(log1p) at *h* = 1, *h* = 7, and *h* = 14, with only weak evidence in favor of ARIMA(log1p) at *h* = 3. Drift also significantly outperforms ETS (log1p) at *h* = 1 and *h* = 14, while the difference at *h* = 7 is not statistically significant. At *h* = 3, there is weak evidence in favor of ETS (log1p) over drift.

The comparison between ETS (log1p) and ARIMA (log1p) is especially informative. At *h* = 1, the two models are statistically indistinguishable under AE loss. At *h* = 3, there is weak evidence favoring ARIMA(log1p), whereas at *h* = 7 and *h* = 14, ETS(log1p) significantly outperforms ARIMA(log1p). These results support the descriptive finding that ARIMA(log1p) is competitive at short horizons, while ETS(log1p) becomes stronger at medium and longer horizons.

Overall, the DM results support three conclusions. First, drift is not merely numerically competitive but significantly outperforms the transformed statistical models at several horizons. Second, ETS (log1p) and ARIMA (log1p) are not interchangeable: their relative performance changes systematically with horizon. Third, differences among the stronger model-based competitors are smaller and less stable at short horizons than at medium and long horizons, underscoring the importance of horizon-specific evaluation.

### Prophet uncertainty behavior

Although Prophet(log1p) is not competitive as a point forecaster in this benchmark, it provides a useful contrast for predictive uncertainty. Table 6 and Fig 9 summarize its horizon-wise point accuracy together with the empirical coverage and width of the nominal 80% prediction intervals.

**Table 6.** Prophet point accuracy and interval summary. Point accuracy is reported using MAE on the original incidence scale. Prediction interval summaries correspond to the nominal 80% interval.

| Horizon | MAE | Coverage80 | MeanWidth80 |
| --- | --- | --- | --- |
| 1 | 50307.81 | 0.9664 | 428686.13 |
| 3 | 56478.30 | 0.9748 | 458445.56 |
| 7 | 72221.35 | 0.9076 | 539173.30 |
| 14 | 118411.71 | 0.8067 | 725819.64 |

**Fig 9.**
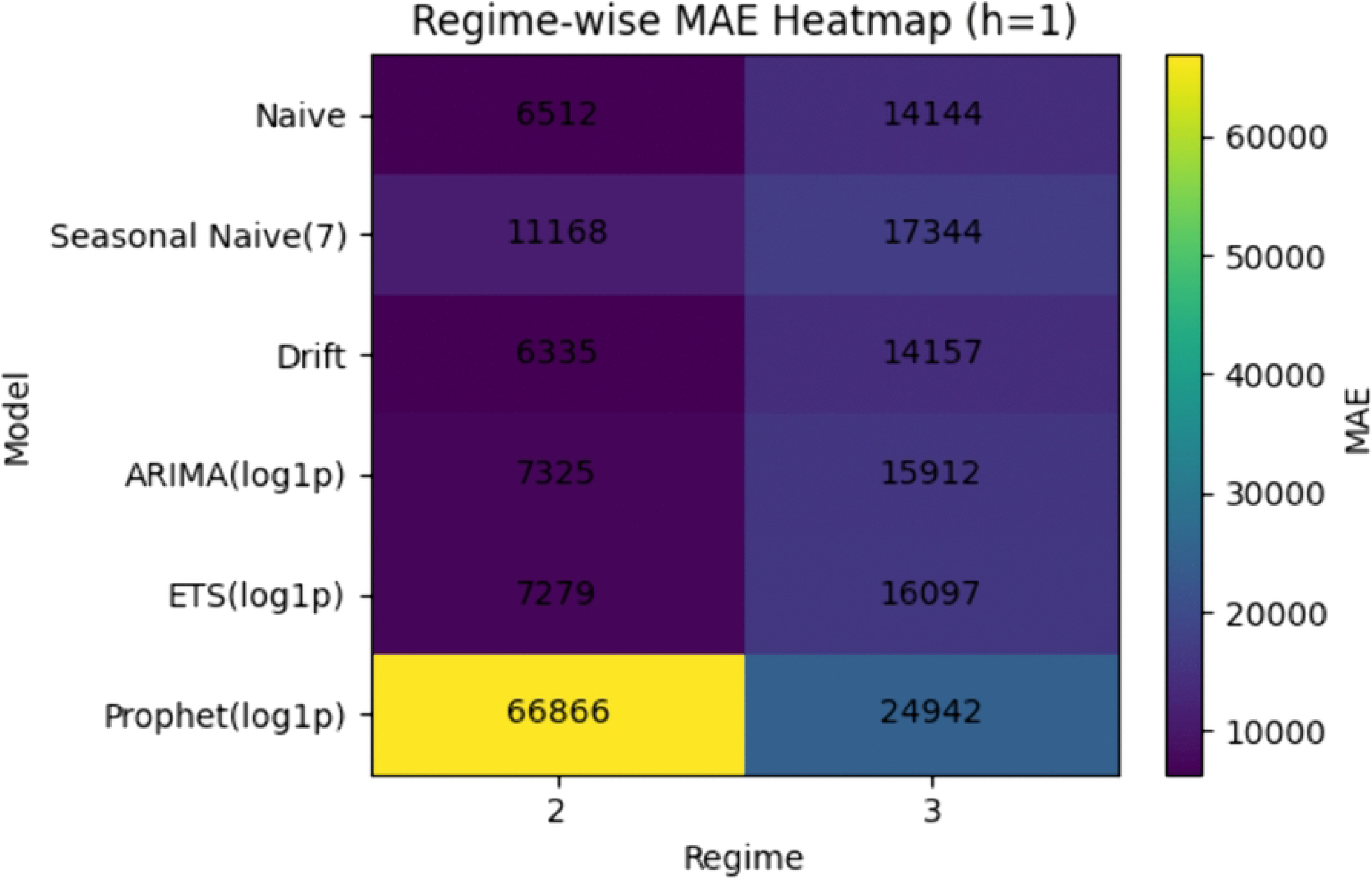
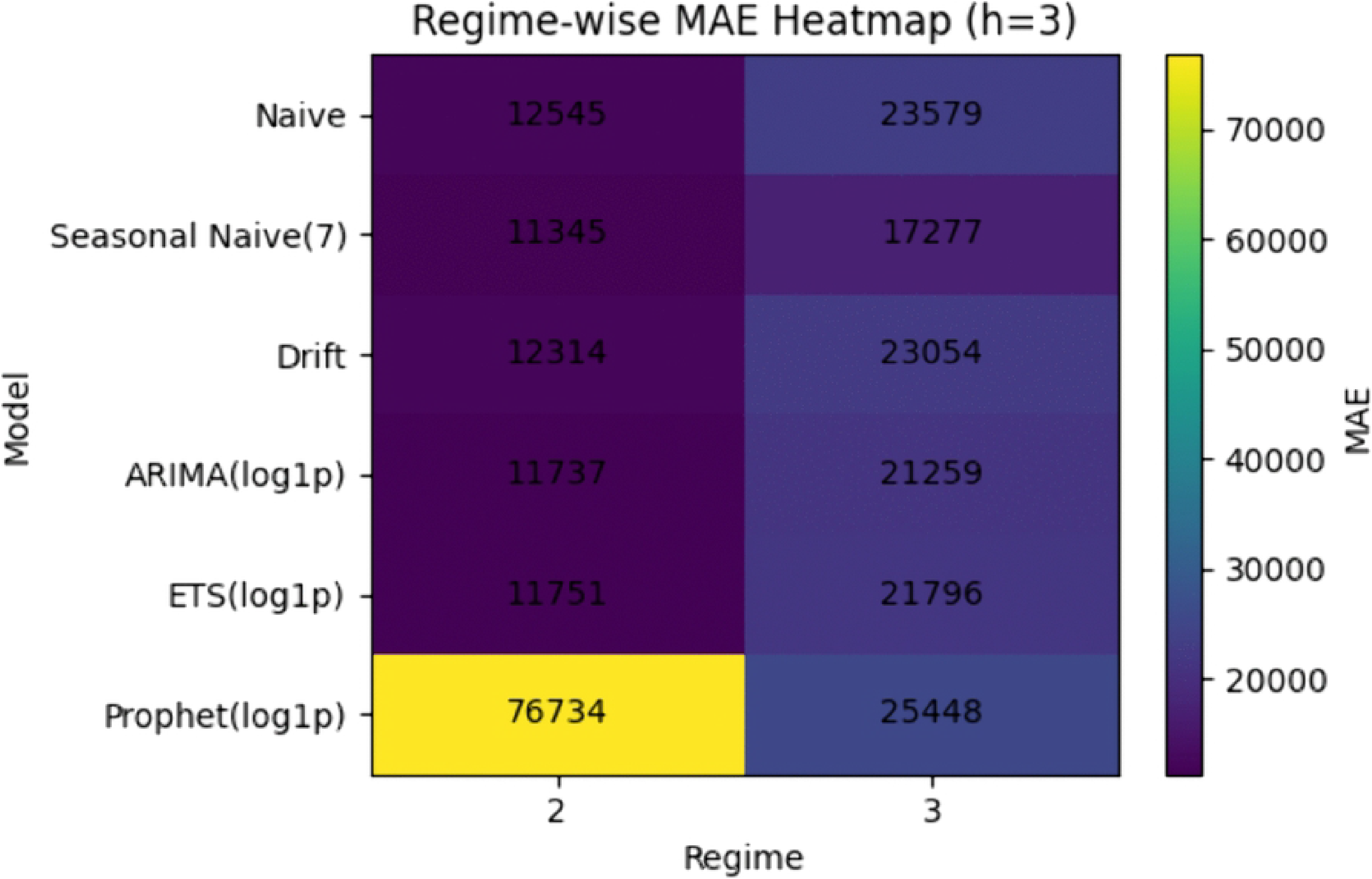
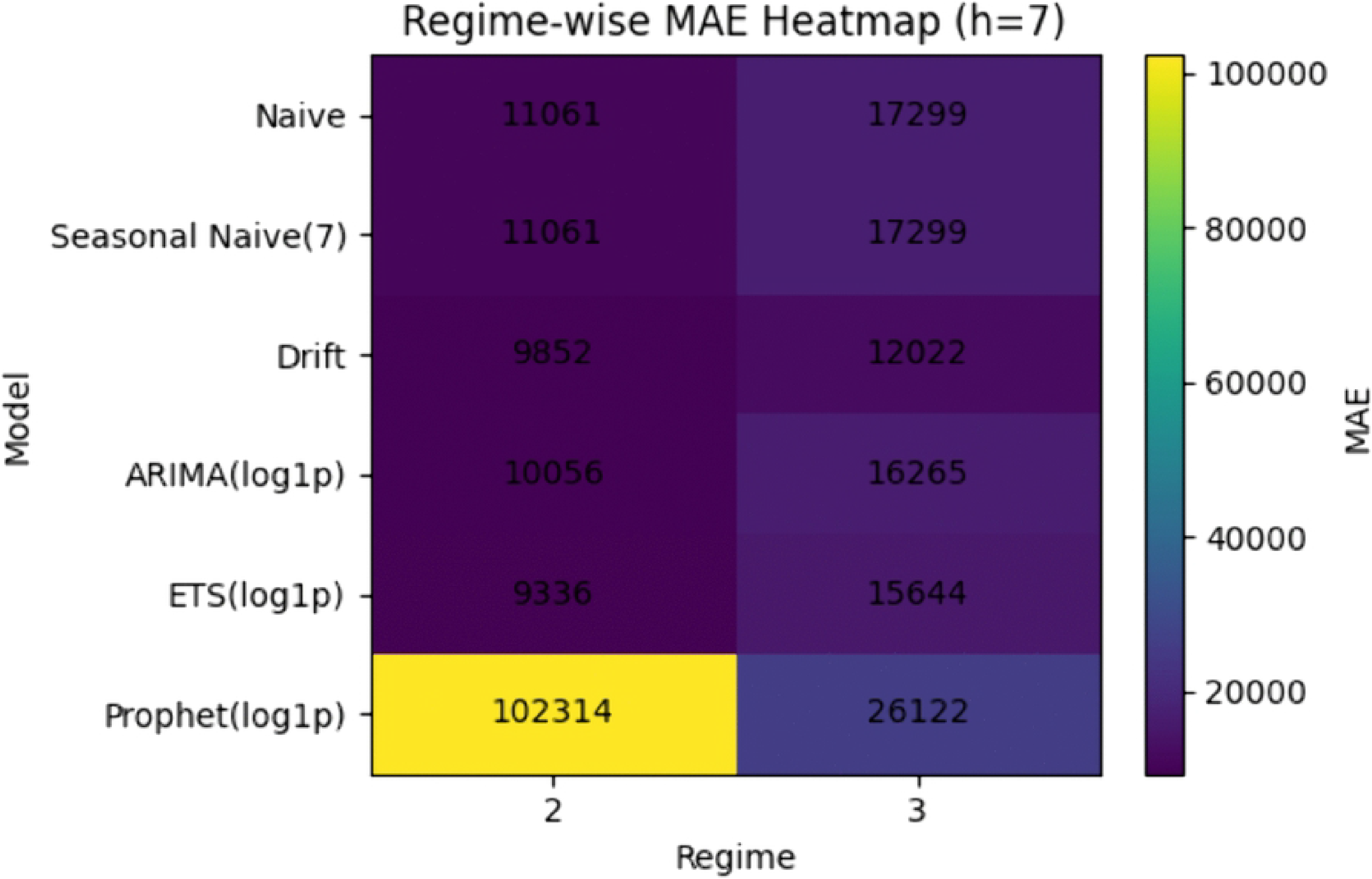
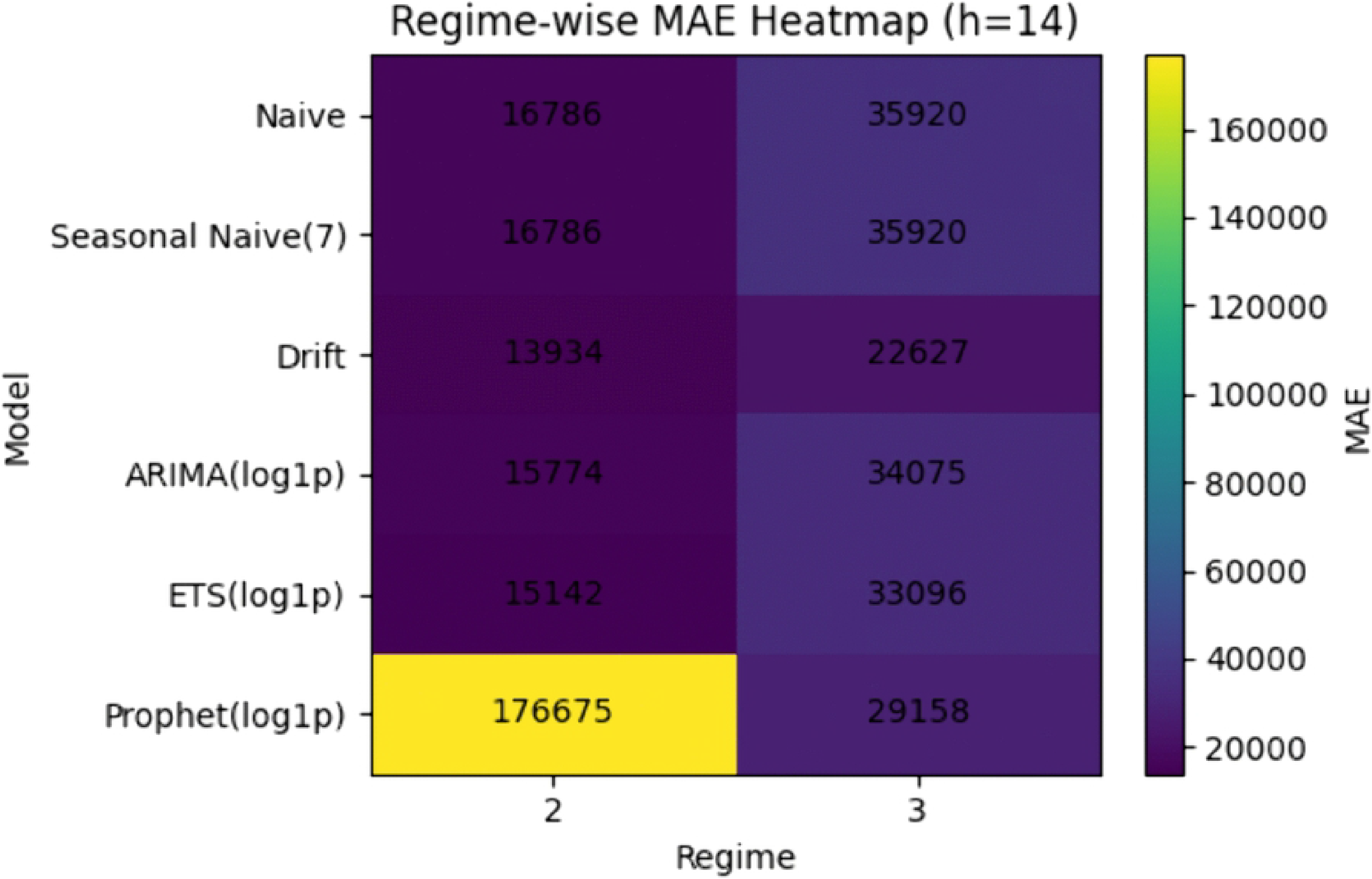
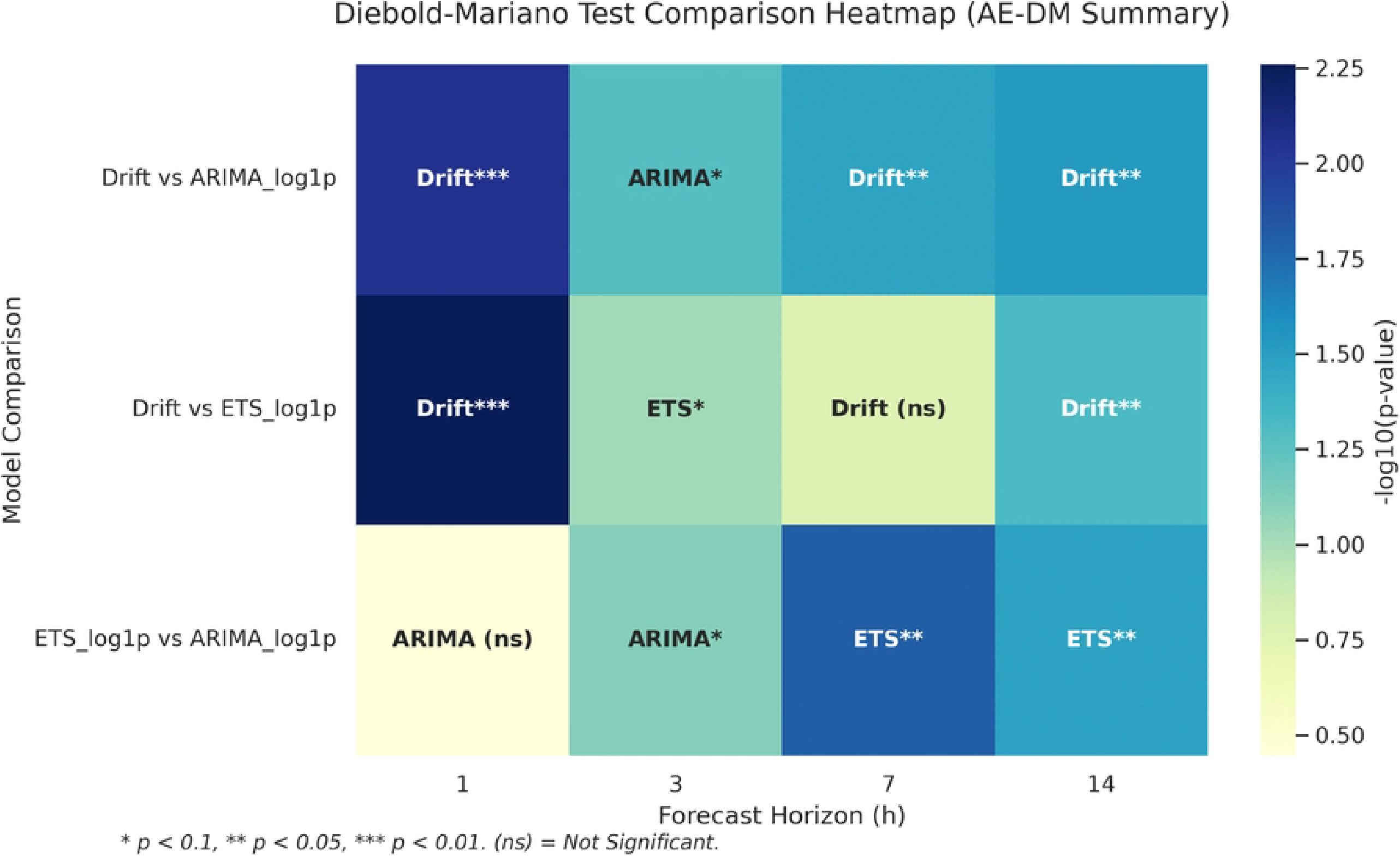
Prophet uncertainty behavior across forecast horizons. Left panel: empirical coverage of the nominal 80% prediction interval. Right panel: interval width as a measure of sharpness. Coverage is well above nominal at short horizons, but this is accompanied by extremely wide intervals, indicating over-conservative uncertainty quantification.

From the perspective of point forecasting, Prophet(log1p) performs poorly at all horizons. Its MAE increases from 50,307.81 at *h* = 1 to 118,411.71 at *h* = 14, far exceeding the corresponding errors of the other benchmark models. Under the lightweight specification used here, Prophet is therefore not competitive for point prediction in this global early-pandemic incidence series.

Its interval behavior is more nuanced, but not necessarily favorable. Empirical coverage is well above the nominal 80% target at short horizons, reaching 0.966 at *h* = 1 and 0.975 at *h* = 3. Coverage then declines with the horizon, falling to 0.908 at *h* = 7 and approaching nominal at *h* = 14 (0.807). Considered in isolation, such high short-horizon coverage might appear attractive; however, it must be interpreted jointly with interval width.

The corresponding prediction intervals are extremely wide. Mean 80% interval width increases monotonically from approximately 428,686 at *h* = 1 to 725,820 at *h* = 14, while median widths are also very large throughout. These values are large relative to the scale of daily incidence over much of the sample, indicating that Prophet attains high short-horizon coverage primarily by producing broad uncertainty bands rather than sharp predictions.

Taken together, these results suggest that Prophet (log1p) is over-conservative in this setting: it combines weak point accuracy with high nominal coverage at short horizons, but only by using excessively wide predictive intervals. Its role in the present benchmark is therefore primarily diagnostic rather than competitive, illustrating that nominal coverage alone is not sufficient for useful probabilistic forecasting when sharpness is poor.

### Robustness analyses

#### Breakpoint stability and regime-label robustness

Because regime segmentation is used only for retrospective stratification, its role is interpretive rather than predictive. It is nevertheless important to assess whether the identified structural phases are sensitive to reasonable changes in segmentation settings. To this end, we vary both the number of breakpoints *K* and the minimum segment length *L*_min_, and re-estimate the retrospective segmentation on the variance-stabilized series *z*_*t*_ = log(1 + *y*_*t*_).

Table 7 and Fig 10 show that the detected breakpoints are moderately stable across settings. A mid-March transition appears in all configurations, although its exact date varies slightly. When more flexible settings are allowed, the early surge is split more finely, producing additional breaks in late February, late March, or early June. By contrast, the baseline setting (*K* = 2, *L*_min_ = 21) yields a simpler segmentation with two broad transitions, on 13 March 2020 and 28 May 2020.

**Table 7.** Breakpoint stability across segmentation settings. Breakpoints are estimated on the variance-stabilized series *z*_*t*_ = log(1 + *y*_*t*_) under alternative choices of the number of breakpoints *K* and minimum segment length *L*_min_.

| Setting | $K$ | $L_{\min}$ | No. of breaks | Detected break dates |
| --- | --- | --- | --- | --- |
| K1_min21 | 1 | 21 | 1 | 2020-03-16 |
| K2_min14 | 2 | 14 | 2 | 2020-03-11, 2020-03-25 |
| K2_min21 | 2 | 21 | 2 | 2020-03-13, 2020-05-28 |
| K3_min21 | 3 | 21 | 3 | 2020-02-29, 2020-03-21, 2020-06-10 |

**Fig 10.**
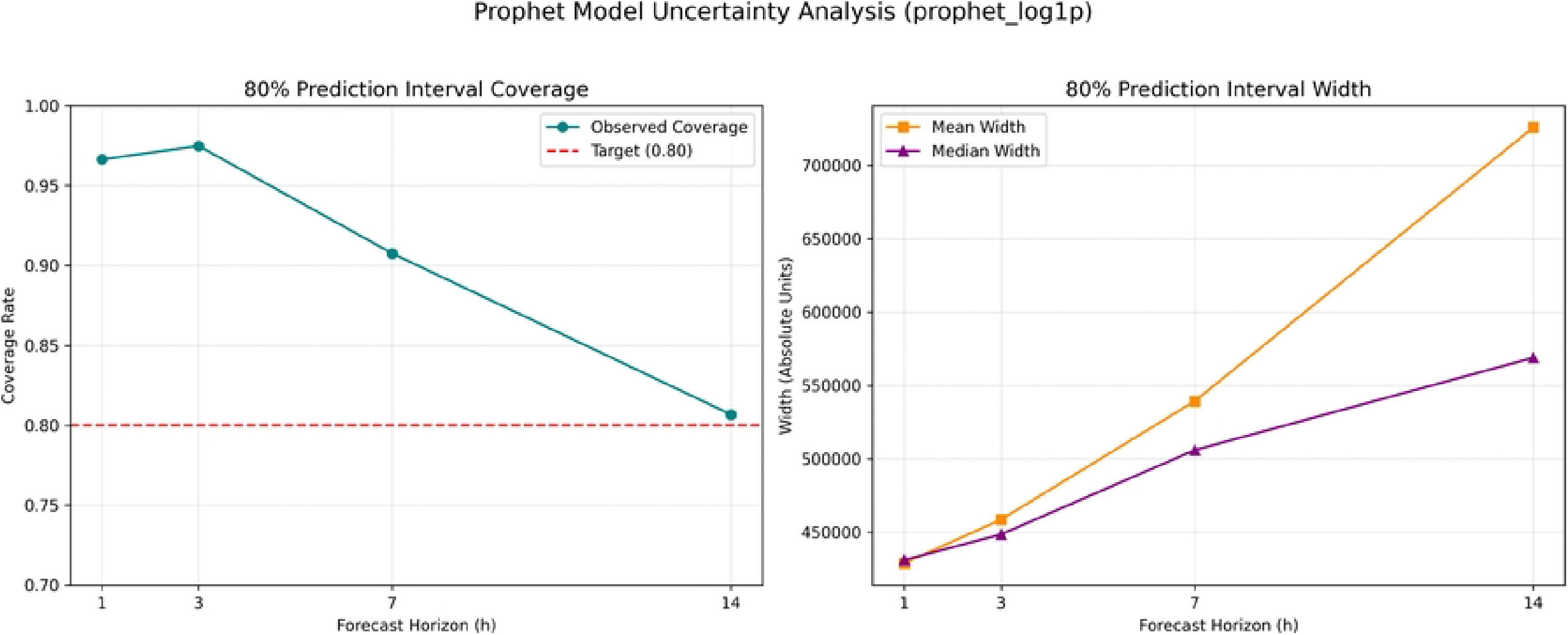
Breakpoint stability across segmentation settings. A mid-March transition is detected consistently across all settings, while more flexible configurations produce additional breaks that split the early surge and later high-incidence period more finely.

This pattern indicates that the observed global incidence series contains a robust major transition during the rapid early-pandemic escalation, together with finer-scale changes whose detection depends on segmentation flexibility. Importantly, these differences in breakpoint placement do not alter the role of segmentation in the paper: regime labels remain purely retrospective and are not used for model estimation or forecast generation.

The qualitative comparison between ETS(log1p) and the naive benchmark is also stable across alternative regime definitions. Across all tested settings, naive remains stronger at the 1-day horizon, whereas ETS(log1p) improves on naive at the 3-, 7-, and 14-day horizons. Thus, although the exact segmentation boundaries are not unique, the horizon-wise interpretation of forecast performance remains broadly unchanged.

#### Training-window sensitivity

Training-window policy can materially affect forecast performance under structural change because shorter windows adapt more quickly to recent shifts but may increase estimation variance, whereas longer or expanding windows provide greater stability at the cost of slower adaptation. To examine this trade-off, we compare ETS(log1p) under four training-window policies: expanding estimation and sliding windows of length 56, 84, and 112 days. The primary comparison uses a common set of forecast origins so that all window policies are evaluated on the same prediction tasks.

Table 8 and Fig 11 show that training-window choice has a substantial effect on ETS(log1p) performance. At the 1-day horizon, naive remains the strongest benchmark under all window settings, and none of the ETS variants surpass it. At longer horizons, however, sliding windows improve substantially on the expanding-window specification.

**Table 8.** Training-window sensitivity for ETS(log1p): common-origin comparison. All window policies are evaluated on the same set of forecast origins. Lower values indicate better point-forecast accuracy.

| Training window | Model | $h = 1$ | $h = 3$ | $h = 7$ | $h = 14$ |
| --- | --- | --- | --- | --- | --- |
| Expanding | ETS(log1p) | 14136.14 | 20302.21 | 14711.68 | 30249.74 |
| Expanding | Naive | 12329.60 | 21597.75 | 16212.92 | 32566.56 |
| Sliding 56 | ETS(log1p) | 13895.01 | 16693.89 | 16070.20 | 26309.60 |
| Sliding 56 | Naive | 12329.60 | 21597.75 | 16212.92 | 32566.56 |
| Sliding 84 | ETS(log1p) | 13474.33 | 18399.52 | 13662.02 | 26662.17 |
| Sliding 84 | Naive | 12329.60 | 21597.75 | 16212.92 | 32566.56 |
| Sliding 112 | ETS(log1p) | 13050.72 | 19199.71 | 14211.95 | 27243.40 |
| Sliding 112 | Naive | 12329.60 | 21597.75 | 16212.92 | 32566.56 |

**Fig 11.**
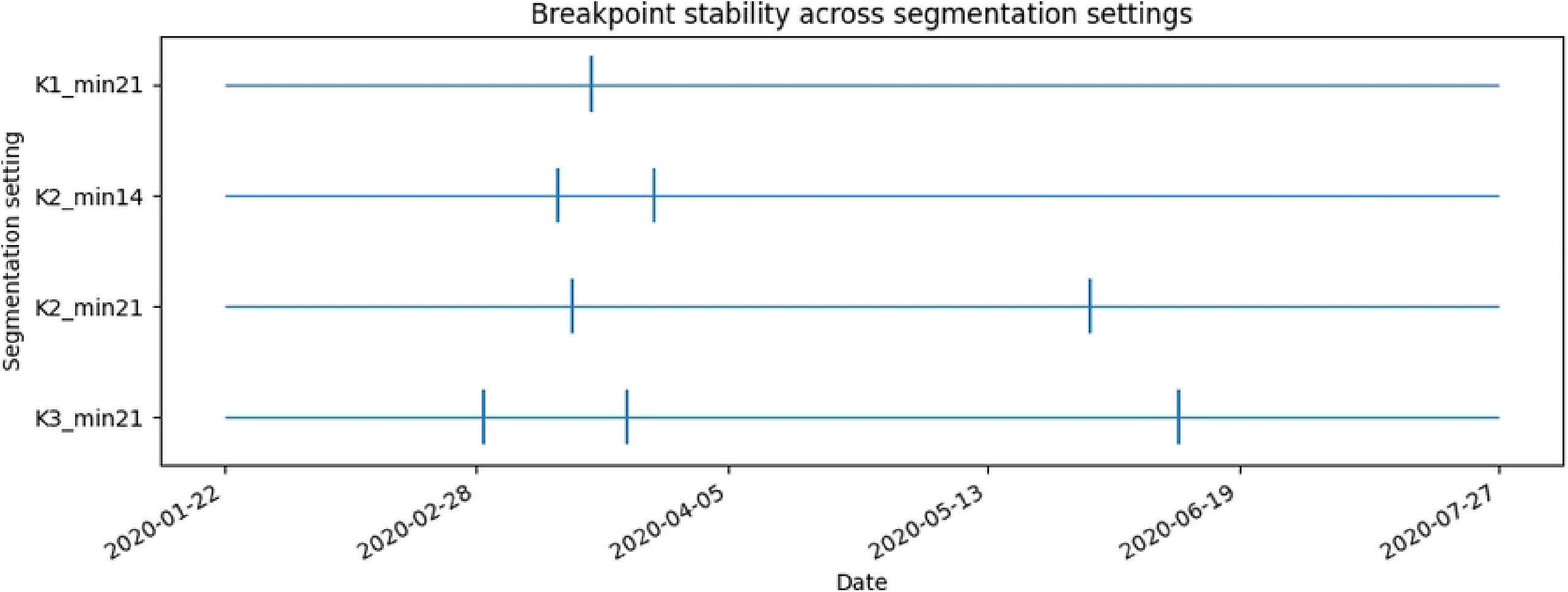
ETS training-window sensitivity under a common-origin comparison. Sliding-window estimation improves ETS(log1p) at medium and long horizons relative to the expanding-window specification, although a very short 56-day window loses stability at *h* = 7. Naive remains best at the 1-day horizon.

The 56-day sliding window achieves the smallest ETS MAE at *h* = 3, indicating that aggressive adaptation can be beneficial at short-to-medium horizons when the signal changes rapidly. This advantage does not persist at *h* = 7, where the 56-day window becomes the weakest ETS variant, suggesting that very short windows can overreact to local fluctuations and reduce stability. By contrast, the 84-day and 112-day windows provide a more balanced compromise. Both improve on the expanding-window specification at *h* = 3, *h* = 7, and *h* = 14, with the 84-day window performing best at *h* = 7 and the 112-day window remaining competitive throughout.

The improvement at longer horizons is especially notable. Relative to the expanding-window specification, all three sliding-window variants reduce MAE at *h* = 14, with the strongest gains obtained under the 56-day and 84-day settings. This pattern supports an adaptation-stability trade-off: moderate sliding windows can improve responsiveness to evolving dynamics without sacrificing too much estimation stability, whereas overly short windows may become unstable at some horizons.

Overall, the common-origin comparison confirms that the training-window policy is an important design choice in nonstationary epidemic forecasting. In this benchmark, naive remains strongest at 1 day, whereas ETS(log1p) benefits from sliding-window estimation at medium and long horizons, particularly when window length balances adaptation and robustness.

#### Coverage-stabilized subsample analysis

A limitation of the full-sample global incidence series is that the number of reporting countries increases monotonically during the early part of the study period. This raises the possibility that some apparent structural changes in forecast performance reflect evolving coverage as well as changes in epidemic dynamics. To assess this issue, we repeat the benchmark on a coverage-stabilized subsample beginning at the first date when the coverage proxy reaches 180 countries (2 April 2020). This cutoff is used as the primary robustness analysis because it preserves a reasonably long series (*n* = 117) while excluding much of the early reporting-expansion phase. Additional cutoffs at 185 and 187 countries are reported as supporting sensitivity checks.

Table 9 and Fig 12 show that trimming the sample changes the short-horizon ranking but does not overturn the broader horizon-dependent pattern. At *h* = 1, naive attains the lowest MAE, with drift close behind and ARIMA(log1p) and ETS(log1p) slightly worse. At *h* = 3, ETS(log1p) becomes the strongest model, outperforming ARIMA(log1p), seasonal naive, drift, and naive. This differs from the full-sample benchmark, where the seasonal naive ranked first at the same horizon.

**Table 9.** Coverage-stabilized rolling-origin benchmark (Subsample ≥ 180 countries). The subsample begins on 2 April 2020 and retains the default minimum training length of 56 days. Lower values indicate better point-forecast accuracy.

| Model | $h = 1$ | $h = 3$ | $h = 7$ | $h = 14$ |
| --- | --- | --- | --- | --- |
| Naive | 14197.08 | 23821.81 | 17288.25 | 35825.90 |
| Seasonal Naive(7) | 17251.27 | 17594.90 | 17288.25 | 35825.90 |
| Drift | 14251.27 | 23478.81 | 12599.71 | 24183.03 |
| ARIMA(log1p) | 14909.77 | 18104.47 | 25049.66 | 53732.93 |
| ETS(log1p) | 15084.11 | 16166.12 | 13884.56 | 21063.48 |

(b) Root Mean Squared Error (RMSE)
| Model | $h = 1$ | $h = 3$ | $h = 7$ | $h = 14$ |
| --- | --- | --- | --- | --- |
| Naive | 18060.57 | 28461.39 | 20759.16 | 39110.95 |
| Seasonal Naive(7) | 20901.20 | 21195.78 | 20759.16 | 39110.95 |
| Drift | 18069.20 | 28271.29 | 16178.47 | 27865.84 |
| ARIMA(log1p) | 17642.39 | 23466.00 | 30912.18 | 62101.41 |
| ETS(log1p) | 18365.92 | 20144.19 | 17872.14 | 26565.15 |

**Fig 12.**
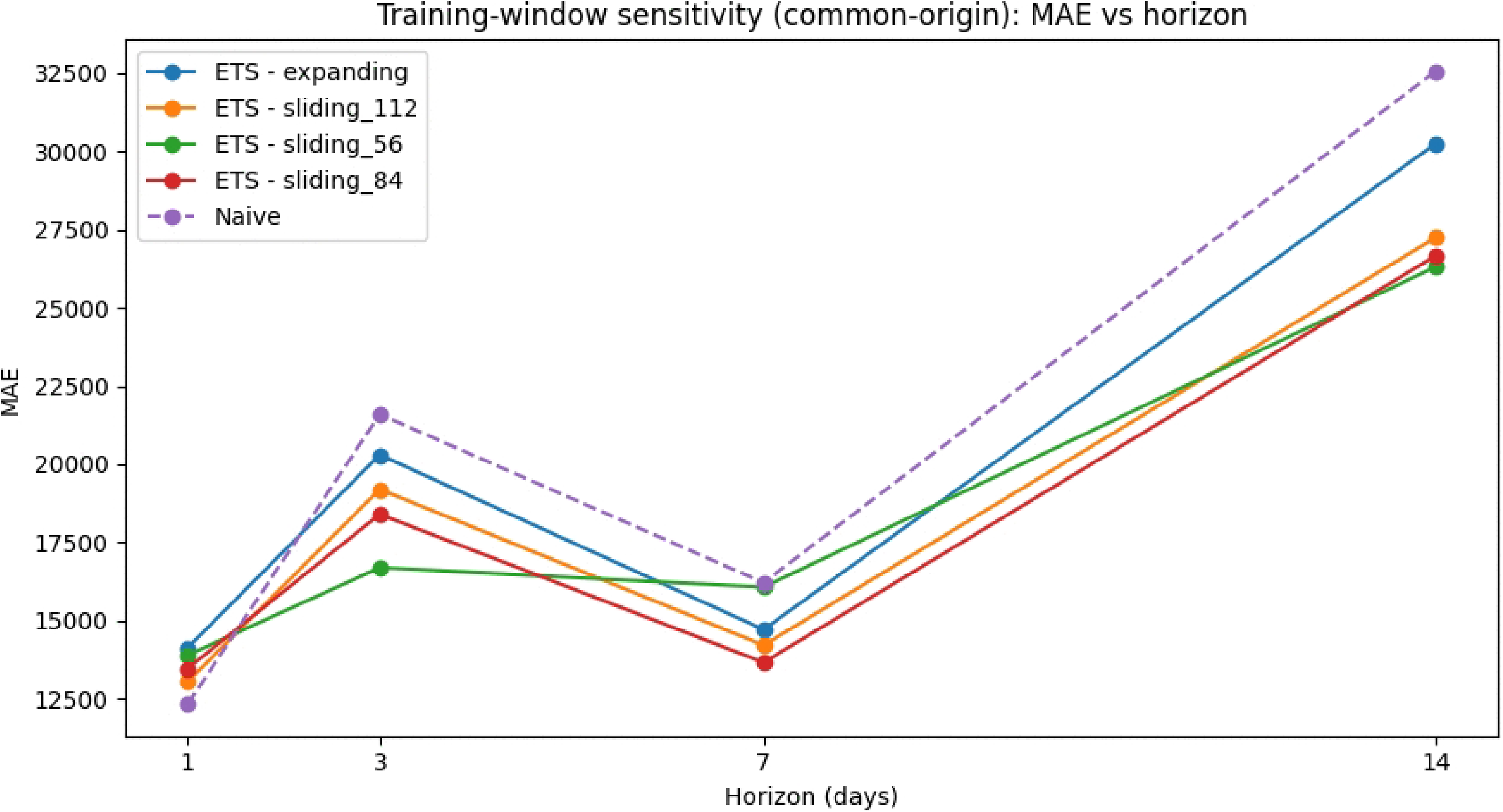
Full-sample versus coverage-stabilized benchmark comparison. Horizon-wise MAE curves for the main coverage-stabilized subsample (≥ 180 countries) show that trimming the early coverage-expansion phase changes the short-horizon ranking, with ETS(log1p) improving at 3 days, while drift remains highly competitive at longer horizons.

At longer horizons, drift remains highly competitive. It achieves the lowest MAE at *h* = 7, while ETS(log1p) is strongest at *h* = 14. Thus, after restricting attention to a period of near-stable reporting coverage, the benchmark shifts toward a clearer division of labor across horizons: naive is strongest at 1 day, ETS(log1p) performs best at 3 and 14 days, and drift remains strongest at 7 days.

Two conclusions follow. First, short-horizon model ranking is sensitive to sample definition, indicating that early coverage expansion influences part of the full-sample benchmark. Second, the main qualitative message survives: forecast ranking remains strongly horizon-dependent, and simple baselines—especially drift—continue to be difficult to outperform consistently even after the reporting footprint stabilizes.

The additional cutoff analyses at 185 and 187 countries yield the same broad pattern. In those later subsamples, ARIMA(log1p) improves relative to the full sample at the 1-day horizon, ETS(log1p) remains competitive at intermediate horizons, and drift continues to perform strongly at 7 and 14 days. These supporting results reinforce the view that the benchmark conclusions are not driven solely by the earliest phase of rapidly expanding coverage.

#### Target-definition robustness

To assess whether the benchmark conclusions depend on the precise construction of the incidence target, we repeat the full rolling-origin analysis using two alternative definitions: the reported daily incidence series (`New cases`) and a reconstructed incidence series obtained as first differences of cumulative confirmed counts, Δ`Confirmed`. This robustness check is motivated by the fact that the two target definitions differ on a limited subset of dates, although they coincide on most of the sample.

The results show that absolute error levels shift modestly under the alternative target construction, but the qualitative ranking pattern remains unchanged. Table 10 and Fig 13 show that drift remains the strongest model at horizons *h* = 1, *h* = 7, and *h* = 14, while seasonal naive remains best at *h* = 3. Likewise, ETS(log1p) continues to outperform ARIMA(log1p) at *h* = 7 and *h* = 14, whereas ARIMA(log1p) remains slightly better at the shorter horizons.

**Table 10.** Cross-target MAE comparison under alternative incidence definitions. The reported target uses `New cases`; the reconstructed target uses first differences of cumulative confirmed counts. Lower values indicate better point-forecast accuracy.

| Model | Target definition | $h = 1$ | $h = 3$ | $h = 7$ | $h = 14$ |
| --- | --- | --- | --- | --- | --- |
| ARIMA(log1p) | Reconstructed | 10572.87 | 15235.62 | 11342.30 | 21380.82 |
| ARIMA(log1p) | Reported | 10716.11 | 15497.77 | 12508.35 | 23002.28 |
| Drift | Reconstructed | 9347.10 | 16429.94 | 10603.13 | 17092.87 |
| Drift | Reported | 9424.21 | 16555.44 | 10708.80 | 17367.38 |
| ETS(log1p) | Reconstructed | 10819.72 | 15294.30 | 11212.61 | 21309.12 |
| ETS(log1p) | Reported | 10761.58 | 15718.33 | 11827.27 | 22233.07 |
| Naive | Reconstructed | 9422.22 | 16775.63 | 13318.76 | 24058.76 |
| Naive | Reported | 9526.42 | 16903.04 | 13524.58 | 24342.65 |
| Seasonal Naive(7) | Reconstructed | 13515.94 | 13568.97 | 13318.76 | 24058.76 |
| Seasonal Naive(7) | Reported | 13607.61 | 13687.83 | 13524.58 | 24342.65 |

**Fig 13.**
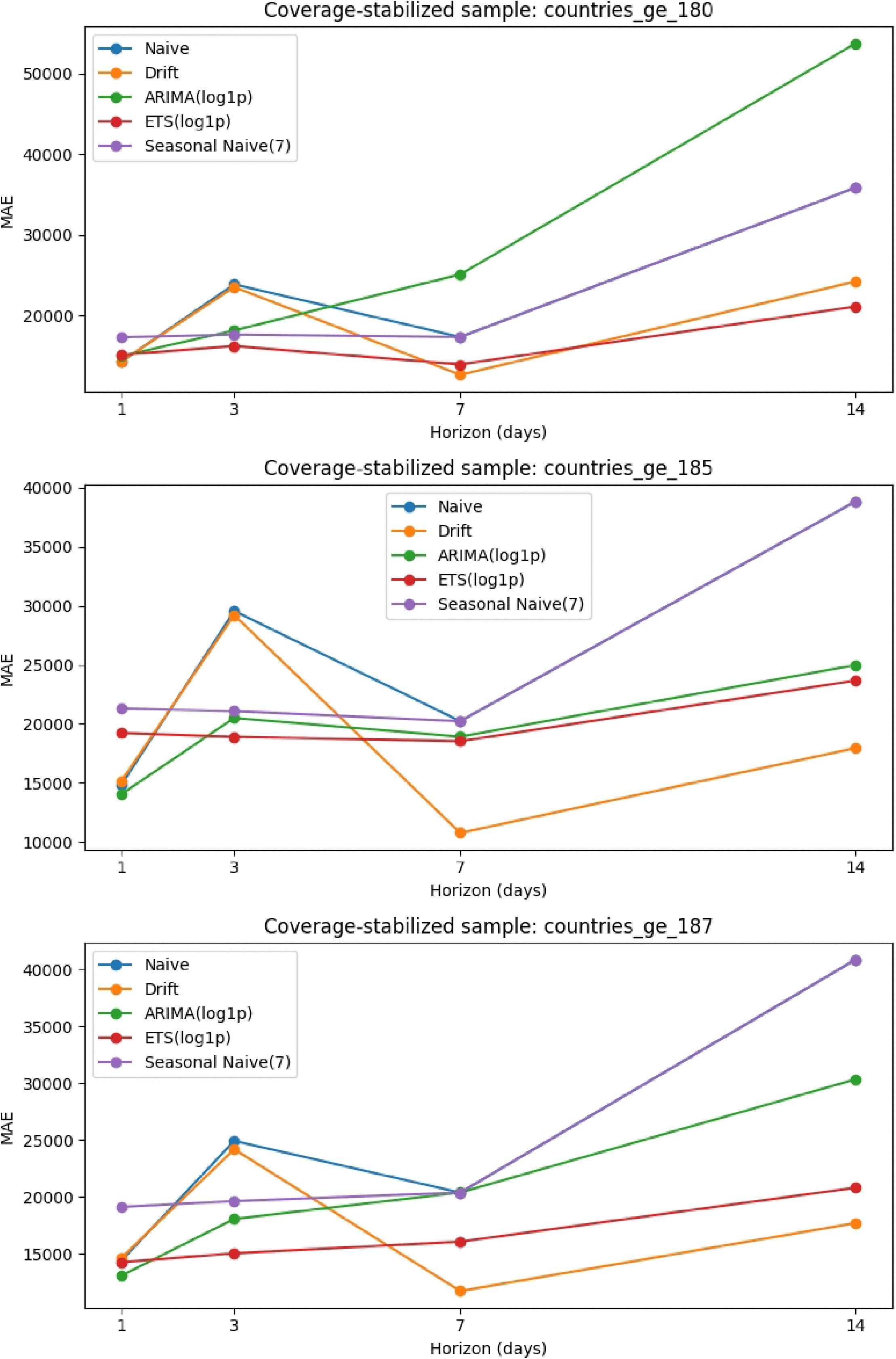
MAE versus horizon under alternative target definitions. Absolute error levels shift modestly between the reported and reconstructed incidence targets, but the horizon-specific ranking pattern remains effectively unchanged.

In general, the reconstructed target yields somewhat lower MAE values for most models, especially at the longer horizons. The reduction is most pronounced for ARIMA(log1p) and ETS(log1p) at *h* = 7 and *h* = 14, but these changes do not alter model ordering. This indicates that the main benchmark conclusions are not driven by the limited discrepancies between the reported and reconstructed daily incidence series.

Overall, the target-definition analysis strengthens the empirical credibility of the study. Although the exact magnitude of forecast error depends somewhat on how daily incidence is constructed, the horizon-specific ranking of models is robust to this alternative target definition.

## Discussion

### Principal findings

This study benchmarked statistical models for forecasting global daily COVID-19 incidence using a rolling-origin evaluation framework suited to nonstationary epidemic conditions. The results show that forecast ranking is strongly horizon-dependent and cannot be summarized by a single overall winner. In the full-sample benchmark, drift performed best at the 1-, 7-, and 14-day horizons, while seasonal naive performed best at 3 days. Among the transformed statistical models, ARIMA(log1p) and ETS(log1p) were competitive at short horizons, but ETS(log1p) outperformed ARIMA(log1p) at 7 and 14 days. By contrast, Prophet(log1p) was not competitive for point forecasting and achieved high short-horizon coverage only through very wide prediction intervals.

The Diebold-Mariano tests reinforced this picture. Drift significantly outperformed ARIMA(log1p) at 1, 7, and 14 days and significantly outperformed ETS(log1p) at 1 and 14 days. ETS(log1p) and ARIMA(log1p) were statistically indistinguishable at 1 day, but ETS(log1p) significantly outperformed ARIMA(log1p) at 7 and 14 days. These results indicate that forecast comparisons for epidemic time series should be made horizon by horizon rather than through a single pooled summary.

The robustness analyses further strengthened the benchmark. Breakpoint locations were reasonably stable across segmentation settings, and the qualitative comparison between ETS(log1p) and naive remained stable by horizon. Moderate sliding windows improved ETS(log1p) at medium and long horizons, coverage-stabilized subsamples changed some short-horizon rankings without overturning the broader horizon-dependent pattern, and alternative target construction based on cumulative confirmed counts left the ranking structure essentially unchanged.

### Why simple baselines remained highly competitive

One of the clearest findings of the study is that simple baselines remained difficult to outperform. This is noteworthy because naive and drift models are often treated mainly as reference points to be surpassed by more structured approaches. Here, however, drift was frequently the strongest model, and naive persistence remained highly competitive at the 1-day horizon.

A plausible explanation lies in the structure of the series itself. The global aggregate smooths across local outbreaks and reporting systems, producing a dominant large-scale trend with persistent level changes. In such a setting, a drift forecast can capture the prevailing direction of change effectively, especially at longer horizons, while naive persistence benefits from short-term continuity. This helps explain why even reasonably well-specified transformed statistical models did not consistently outperform the simpler baselines.

The strong showing of seasonal naive at the 3-day horizon is also informative. It suggests that weekly reporting regularities remain detectable even in an aggregate global series, although not strongly enough to dominate across all horizons. After trimming to coverage-stabilized subsamples, this advantage became less pronounced, indicating that part of the short-horizon seasonal signal may be linked to the transitional structure of the early reporting period.

### Implications of structural change and coverage stabilization

The regime-stratified results and coverage-stabilized subsample analyses provide a more nuanced view of the benchmark. Retrospective segmentation identified clear structural changes in the observed series, but these should not be interpreted as purely epidemiological regimes. Because the data are global aggregates and reporting coverage expanded substantially during the early sample, the detected regimes reflect changes in the observed process rather than clean causal transitions in disease transmission.

This distinction matters. When the benchmark was repeated on the coverage-stabilized subsample beginning at 180 countries, short-horizon rankings changed: naive performed best at 1 day, ETS(log1p) at 3 and 14 days, and drift at 7 days. Thus, part of the short-horizon full-sample ranking depends on the early expansion of the reporting footprint. At the same time, the broader conclusion remained intact: forecast ranking was still strongly horizon-dependent, and simple baselines-especially drift-continued to be highly competitive after coverage stabilized.

These findings suggest that the main results are not merely artifacts of the earliest and most unstable part of the dataset. Instead, they reflect a persistent interaction among forecast horizon, model structure, and the evolving characteristics of the observed series.

### Training-window policy and the adaptation-stability trade-off

The training-window analysis revealed a clear trade-off between adaptation and stability. Expanding windows preserve all available history and therefore stabilize parameter estimation, but they can adapt too slowly when the data-generating process changes rapidly. Sliding windows adapt more quickly by discarding older observations, but they may increase estimation variance when the window becomes too short.

The results were consistent with this logic. For ETS(log1p), moderate sliding windows improved performance at medium and long horizons relative to the expanding-window benchmark. The 56-day window performed best at 3 days but deteriorated at 7 days, whereas the 84-day and 112-day windows provided a better balance between responsiveness and stability. Under epidemic nonstationarity, the training-window policy can therefore materially influence model ranking and should be treated as a substantive design choice rather than a minor implementation detail.

### Prophet and the distinction between calibration and usefulness

Prophet(log1p) illustrates that high nominal coverage does not necessarily imply useful uncertainty quantification. In this study, Prophet achieved high short-horizon coverage, but its prediction intervals were extremely wide and its point forecasts were poor. In other words, the model was calibrated only in a loose sense because it was insufficiently sharp.

This distinction is important in epidemic forecasting, where decision-makers need uncertainty estimates that are not only statistically plausible but also operationally useful. Broad intervals that cover almost all plausible outcomes may be difficult to interpret or act upon. These results therefore suggest that probabilistic forecast evaluation should consider both calibration and sharpness, rather than coverage alone.

### Public health relevance

These findings have two practical public-health implications. First, horizon-specific model choice matters. A model that performs well at 1 day need not be strongest at 7 or 14 days, and vice versa. This is relevant because different operational tasks correspond to different forecast horizons: very short-horizon forecasts may support daily situational monitoring, whereas 7-to 14-day forecasts are more relevant for planning staffing, bed capacity, logistics, and communication.

Second, simple baselines deserve serious consideration in surveillance settings. Drift and naive models remained highly competitive under structural change and should not be treated merely as trivial comparators. In settings where speed, transparency, and reproducibility matter, such models may provide useful reference forecasts or even strong primary benchmarks against which more elaborate methods should be judged.

More broadly, the results support the use of rolling-origin protocols, horizon-stratified evaluation, and explicit checks for reporting changes in public-health forecasting. This is especially important in epidemic surveillance, where the observed series can change because of both underlying transmission and evolving measurement systems.

### Limitations

This study has several limitations. First, it relies on a single global aggregate incidence series from the early pandemic. Although this makes the benchmark transparent and reproducible, it limits generalizability and smooths important local variation.

Second, the regime segmentation is retrospective and descriptive, not causal or predictive. Third, the benchmark focuses on statistical models and simple baselines rather than mechanistic, ensemble, or more advanced machine-learning approaches. Fourth, the lightweight Prophet specification may not reflect the best achievable Prophet configuration. Finally, despite the robustness checks, the analysis remains dependent on publicly aggregated surveillance data whose reporting conventions may evolve.

### Future work

Several extensions would strengthen this line of research. A natural next step is to repeat the benchmark on multi-country or multi-region panels, where local dynamics and reporting structure can be studied more directly. It would also be valuable to compare simple baselines and transformed statistical models with regime-switching, time-varying parameter, ensemble, or mechanistic approaches. Broader probabilistic evaluation using proper scoring rules, beyond Prophet alone, is another important direction. Finally, incorporating exogenous factors such as mobility, testing intensity, or interventions may help identify when more complex models yield reliable improvements over strong baselines.

## Conclusion

This study evaluated statistical forecasting methods for global daily COVID-19 incidence using a rolling-origin backtesting framework tailored to nonstationary epidemic data. Forecast performance was strongly horizon-dependent: drift performed best at the 1-, 7-, and 14-day horizons, while seasonal naive performed best at 3 days. Among the transformed statistical models, ETS(log1p) and ARIMA(log1p) were consistently competitive, with ETS(log1p) outperforming ARIMA(log1p) at longer horizons. Prophet(log1p) produced poor point forecasts and achieved high nominal coverage only through very wide intervals.

The Diebold–Mariano tests confirmed significant differences in several pairwise comparisons, particularly in favor of drift at short and long horizons and in favor of ETS(log1p) over ARIMA(log1p) at the longer horizons. No single model dominated across all horizons, reinforcing the need for horizon-specific evaluation.

The robustness analyses strengthened the credibility of the results. Breakpoints were reasonably stable, moderate sliding windows improved ETS(log1p) at medium and long horizons, coverage-stabilized subsamples shifted some short-horizon rankings without changing the broader pattern, and alternative target definitions altered absolute error levels only modestly without changing model ordering.

Overall, three lessons emerge. First, epidemic forecasting benchmarks should report horizon-specific results rather than aggregated summaries. Second, simple baselines such as naive and drift can remain highly competitive under structural change and should be treated as serious reference models. Third, evaluation frameworks should account explicitly for reporting and target-construction effects so that forecast comparisons reflect genuine predictive skill rather than artifacts of evolving surveillance data.

Future work could extend this benchmark to multi-country data, broaden probabilistic evaluation, and compare statistical models with mechanistic or ensemble approaches to clarify when more complex methods provide reliable gains over strong baselines.

## Data Availability

The data underlying the results presented in the study are available from https://www.kaggle.com/datasets/imdevskp/corona-virus-report

## Acknowledgements

The authors gratefully acknowledge the academic support of the Pan-African University Institute for Basic Sciences, Technology, and Innovation (PAUSTI).

## Data availability statement

The data used in this study are derived from publicly available COVID-19 surveillance sources cited in the manuscript. The processed time series and code used for the analyses are available from the corresponding author upon reasonable request.

## Author contributions

Michael Marko Sesay: Conceptualization, methodology, data curation, formal analysis, software, visualization, writing-original draft.

Moise Saleh Wembo: validation, writing-review and editing.

All authors read and approved the final manuscript.

